# Separation-like irregularity and sample size optimism in high-discrimination logistic prediction models

**DOI:** 10.64898/2026.01.21.26344587

**Authors:** Zonghao Liu, Ye Liang, Louis Shuo Wang, Jiguang Yu, Jingfeng Liu

**Author notes:** Corresponding authors. (LSW), (JFL). These authors contributed equally to this work as co-first authors (Zonghao Liu, Ye Liang, Louis Shuo Wang, Jiguang Yu).

## Abstract

Closed-form minimum sample size criteria for developing logistic prediction models, such as the Riley framework implemented in `pmsampsize`, are widely used but may become optimistic when anticipated discrimination is high. We conducted a Monte Carlo simulation study to compare the formula-based recommended development sample size, *n*_Riley_, with an empirical required sample size, *n*_*req*_, *defined by out-of-*sample calibration-slope stability under repeated development sampling. Scenarios fixed the candidate parameter dimension at *p* = 10 and crossed predictor distribution (normal, standardized skewed continuous, binary), signal density (dense versus sparse), prevalence (*ϕ* ∈ {0.05, 0.10, 0.20}), and target discrimination (AUC_target_ ∈ {0.70, 0.75, 0.80, 0.85, 0.90}), with intercept and signal strength calibrated to match targets. We defined *n*_req_ as the smallest *n* such that 𝔼 (*b*_*n*_) ≥ 0.90 and Pr(*b*_*n*_ < 0.80) ≤ 0.20, where *b*_*n*_ is the truth-based logit-scale calibration slope evaluated on a large fixed validation covariate set. At moderate discrimination, *n*_Riley_ approximated *n*_req_, but as discrimination increased the formula increasingly underestimated the sample size required for calibration stability, with large deficits at AUC_target_ = 0.90. Separation-like behavior (extreme fitted risks and linear predictors) at *n* = *n*_Riley_ became common in high-discrimination settings despite nominal convergence, providing a plausible mechanism for formula optimism. These findings support augmenting formula-based planning with targeted simulation stress tests and instability diagnostics when high discrimination is anticipated.

## Introduction

Clinical prediction models aim to estimate an individual’s risk of a future or concurrent health outcome as a function of routinely collected patient characteristics, thereby supporting risk stratification and decision-making in contexts such as screening, treatment selection, and perioperative planning [1–3]. For binary outcomes, logistic regression remains a widely used modelling framework because it targets the conditional event probability via the logit link, yields parameters that are interpretable on the log-odds scale, and is straightforward to implement in clinical workflows. Moreover, in many applications with structured, tabular predictors, logistic regression can provide competitive predictive performance relative to more complex algorithms when sample sizes are finite and the signal-to-noise ratio is modest [4].

The clinical utility of a prediction model depends not only on discrimination, but critically on calibration [5]. While discrimination focuses on ranking, Alba et al. [6] caution that a model can distinguish patients well yet still yield clinically misleading risk estimates. Van Calster et al. [7] characterize this frequent oversight as the ‘Achilles heel’ of predictive analytics, warning that systematic over- or under-estimation of risk is common even in models with high AUCs. As Vickers et al. [8] demonstrate, miscalibration distorts decision-analytic measures (Net Benefit), potentially triggering harmful therapeutic errors such as treating low-risk patients or neglecting those at high risk. A convenient summary of calibration for logistic prediction models is the logit-scale calibration model in a validation sample,

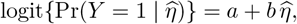

where 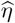 is the fitted linear predictor from the development model, *a* is the calibration intercept (calibration-in-the-large), and *b* is the calibration slope. Values *b* < 1 indicate that predicted log-odds are too extreme (overfitting), whereas values *b* > 1 indicate that predicted log-odds are insufficiently extreme (underfitting or over-regularization) [9, 10]. Because calibration governs absolute risk accuracy, miscalibration can lead to clinically consequential errors even when discrimination is acceptable.

Adequate development sample size is a central prerequisite for achieving stable calibration. When the development dataset is too small relative to the effective complexity of the modeling task, maximum-likelihood coefficient estimates tend to be overly variable and, in repeated sampling, overly extreme in magnitude [11]. This manifests as optimistic apparent performance in the development sample and degraded calibration in new individuals, typically summarized by a calibration slope below one. In this sense, overfitting is naturally expressed on the logit scale as excessive variability of 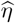 relative to the outcome-generating process. Contemporary sample size criteria for prediction modeling therefore emphasize explicit control of overfitting via shrinkage of the linear predictor. Under standard regularity conditions, the expected global shrinkage of the fitted linear predictor is closely aligned with the expected external calibration slope, providing an interpretable bridge between shrinkage-based planning targets and logit-scale calibration behaviour in new data [12, 13].

Historically, development sample size planning for logistic regression has often relied on simple heuristics based on events-per-variable (EPV), most notably a threshold of at least 10 events per predictor parameter [14]. Subsequent research has demonstrated that Events Per Variable (EPV) heuristics are neither necessary nor sufficient for reliable model development. While acceptable estimation can occur with fewer than 10 events per parameter in some settings [15], substantially larger samples are often required when applying complex modeling techniques such as Support Vector Machines and Neural Networks [16] or when low-prevalence predictors are present [17]. Furthermore, Van Smeden et al. [18] exposed a fundamental flaw in the metric itself, showing that bias can be reduced by increasing the total sample size even if the EPV remains constant.

These limitations have motivated a shift toward performance-based sample size methods. Van Smeden et al. [19] argue that sample size planning should account for the number of predictors, the total sample size and the events fraction to minimize prediction error. In parallel, Riley and colleagues proposed minimum development sample size criteria for multivariable logistic prediction models that target limited overfitting and adequate estimation through three complementary requirements: achieving a prespecified global shrinkage factor, limiting optimism in a measure of model fit via a pseudo-*R*^2^ criterion, and ensuring adequate precision in the estimation of the model intercept [20, 21].

While these criteria have been operationalized for applied use in the pmsampsize R package [22], recent evaluations suggest that the closed-form recommendations may be insufficient under certain design regimes. Notably, Pavlou et al. [23] demonstrate that these approximations systematically underestimate the required sample size in settings where discrimination is high (e.g., C-statistic = 0.85 and 0.9). Furthermore, Pavlou et al. [24] argue that the analytical approximations underpinning the shrinkage and model-fit components may become less accurate in challenging settings, specifically highlighting the need to account for performance variability when determining sample size. This thesis therefore focuses on the required development sample size for binary-outcome prediction models, defined in terms of out-of-sample calibration-slope stability under repeated development sampling, and evaluates where the `pmsampsize` recommendations agree with, or become optimistic relative to, simulation-based requirements.

The overarching aim is to evaluate whether closed-form minimum development sample size recommendations for unpenalized logistic regression remain adequate when anticipated discrimination is high, using an estimand aligned with out-of-sample calibration on the logit scale.

- **Primary objective:** Quantify the discrepancy Δ*n*(AUC_target_, *ϕ*_target_) = *n*_req_ − *n*_Riley_ as a function of target discrimination AUC_target_ (and across prevalence strata *ϕ*_target_), where *n*_req_ is defined by prespecified repeated-sampling stability targets for the out-of-sample logit-scale calibration slope.
- **Secondary objective (robustness):** Assess whether the discrimination–discrepancy relationship is robust to clinically plausible predictor data-generating structures by repeating the primary comparison across (i) alternative predictor distributions (normal, standardized skewed continuous, and binary) and (ii) alternative signal-density configurations (dense versus sparse) while holding the candidate parameter dimension fixed.
- **Mechanistic objective:** At *n* = *n*_Riley_, document the frequency of separation- and convergence-related irregularity in unpenalized logistic regression fits (e.g., extreme fitted risks and separation-warning indicators), and relate these diagnostics to the observed optimism of *n*_Riley_ relative to *n*_req_ in high-discrimination regimes.

This work makes three contributions to performance-based development sample size planning for binary-outcome prediction models:

- **Empirical map of agreement versus optimism of closed-form recommendations**. We provide a systematic, scenario-based characterization of where the Riley/`pmsampsize` minimum sample size recommendation *n*_Riley_ aligns with, versus underestimates, the simulation-defined requirement *n*_req_ when calibration-slope stability under repeated development sampling is used as the planning target, with emphasis on how the discrepancy varies with target discrimination (AUC).
- **A reproducible diagnostic protocol to interrogate regularity in high-discrimination regimes**. We operationalize and report a transparent set of convergence- and separation-related flags (e.g., extreme fitted risks and linear predictor thresholds) evaluated at *n* = *n*_Riley_, enabling investigators to document when likelihood-based approximations that motivate closed-form shrinkage calculations are plausibly strained in practice and to connect these diagnostics to observed optimism.
- **Practical planning and reporting guidance for applied researchers**. We translate the simulation and diagnostic findings into actionable recommendations: using `pmsampsize` as a principled baseline, augmenting it with a targeted stress test when high discrimination is anticipated, and reporting assumptions, calibration summaries, and instability diagnostics in a manner aligned with contemporary expectations for transparent prediction modeling and critical appraisal.

Section Methods describes the simulation study design, including the data-generating mechanisms, calibration of scenario parameters to target prevalence and discrimination, the definition and estimation of the simulation-based required sample size *n*_req_, and the computation of the formula-based benchmark *n*_Riley_ using `pmsampsize`. Section Results presents the primary results on the discrepancy *n*_req_ − *n*_Riley_ as a function of target discrimination, followed by robustness analyses across predictor distributions and signal-density structures and a mechanistic assessment of separation- and convergence-related irregularity at n = *n*_Riley_. Section Applied Illustration provides an applied illustration in a cardiac surgery cohort comparing unpenalized and ridge-penalized logistic regression under bootstrap optimism correction to demonstrate how calibration-focused reporting can be implemented in practice. Section Discussion interprets the findings in relation to existing methodological guidance, outlines practical implications for sample size planning and model development in high-discrimination settings, and discusses limitations and priorities for future work.

## Methods

### Overview of study design

We conducted a Monte Carlo simulation study to evaluate the operating characteristics of the minimum development sample size criteria proposed by Riley and colleagues for binary-outcome prediction models, as implemented in the `pmsampsize` package, the overall simulation workflow is visualized in Fig 1. The study was designed to compare the formula-based recommended sample size, denoted *n*_Riley_ (Subsubsection Formula-based benchmark (*n*_Riley_) in Section Methods), with an empirically determined required development sample size, denoted *n*_req_ (Subsubsection Empirical calibration stability criteria *n*_req_ in Section Methods), defined by prespecified calibration-stability targets under repeated sampling from a known data-generating mechanism (Subsection Simulation scenarios in Section Methods). For each scenario, the data-generating mechanism was fully specified and calibrated to achieve pre-defined population-level discrimination and outcome prevalence, so that discrepancies between *n*_Riley_ and *n*_req_ could be attributed to the performance of the closed-form approximation rather than to mismatch between nominal and achieved design characteristics.

**Fig 1.**
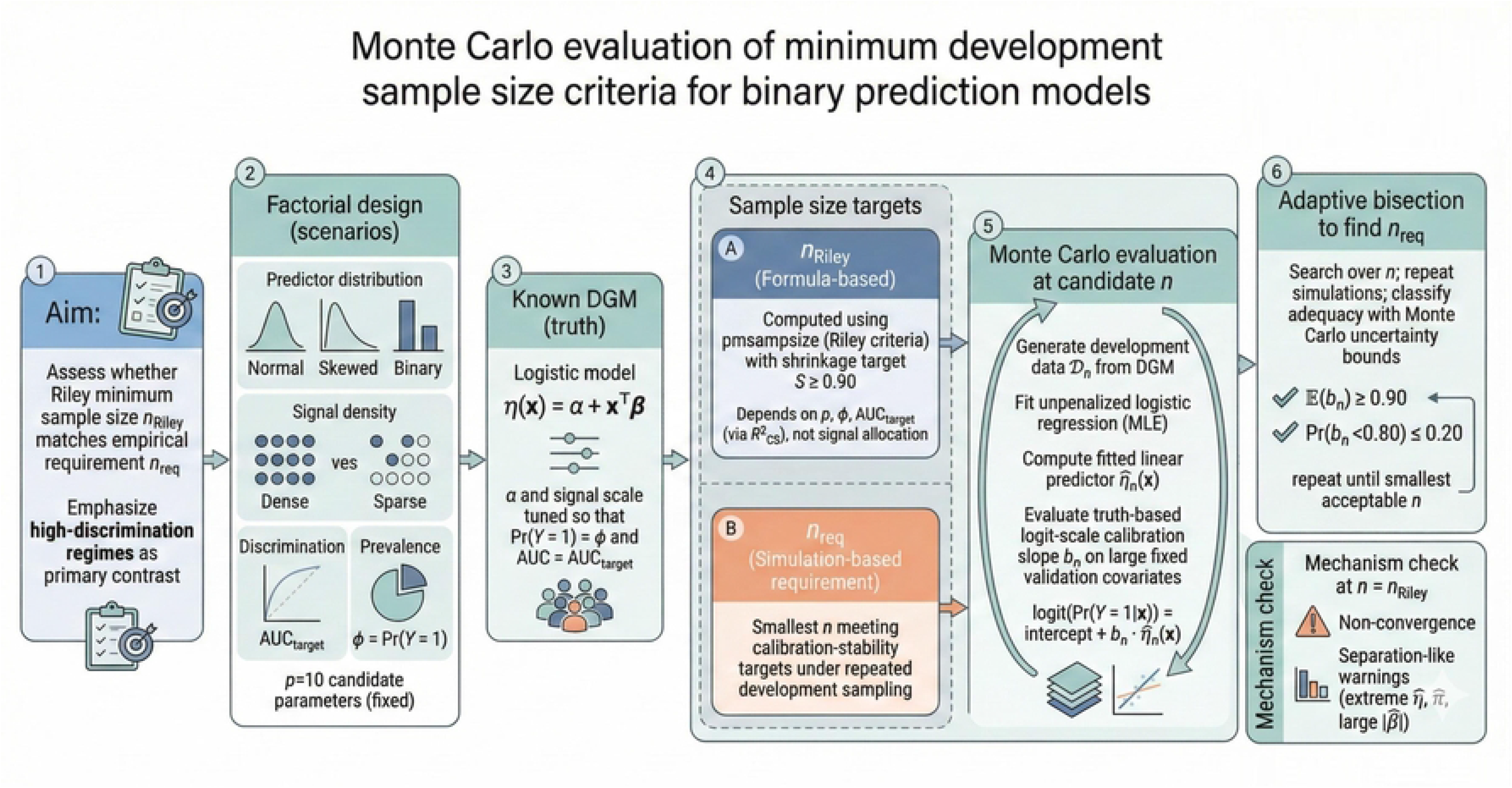
Overall simulation flowchart.

The simulation scenarios were defined by a factorial grid crossing four design factors: predictor distribution, signal density, target discrimination, and prevalence. Predictor distributions were selected to represent common structures encountered in applied prediction modeling, including normal predictors, skewed continuous predictors, and binary predictors (Subsubsection Predictor distributions in Section Methods). Signal density described how prognostic information was distributed across the *p* candidate predictor parameters, contrasting a dense configuration in which all candidate predictors carried signal with a sparse configuration in which signal was concentrated in a subset of predictors while the total number of candidate parameters was held fixed (Subsubsection Signal density in Section Methods). Discrimination was controlled through pre-specified target values of the area under the receiver operating characteristic curve, AUC_target_, and prevalence was controlled through pre-specified target event proportions, *ϕ*_target_. Within each combination of predictor distribution and signal density, the intercept and signal-strength scaling parameter were calibrated so that the induced population event probability satisfied Pr(*Y* = 1) = *ϕ*_target_ and the induced population discrimination satisfied AUC = AUC_target_ (Subsubsection Calibration of scenario parameters to design targets in Section Methods).

Although the factorial grid included multiple prevalence strata, the primary scientific contrast in this paper concerns discrimination. Specifically, we evaluated how the discrepancy between the formula-based recommendation and the empirically required sample size varies as discrimination increases, with emphasis on high-discrimination regimes. For each scenario in the grid, we computed *n*_Riley_ using `pmsampsize` under a fixed global shrinkage target, and we estimated *n*_req_ by an adaptive bisection search over candidate development sample sizes, where each candidate *n* was evaluated by repeated simulation and truth-based logit-scale calibration assessment on a large fixed validation covariate set. In addition, to investigate whether numerical irregularities could plausibly contribute to discrepancies in high-discrimination settings, we recorded pre-specified convergence and separation-related diagnostics for unpenalized logistic regression fits at *n* = *n*_Riley_ and summarized their empirical frequencies across replicates (Subsubsection Separation and convergence diagnostics in Section Methods).

### Notation and estimands

Let **X** ∈ ℝ^*p*^ denote the vector of *p* candidate predictor variables and let *Y* ∈ {0, 1} denote the binary outcome. For each scenario, outcomes are generated under a correctly specified logistic regression model with a linear predictor

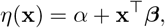

where *α* ∈ ℝ is the intercept and ***β*** ∈ ℝ^*p*^ is the vector of slope coefficients. The corresponding conditional event probability is

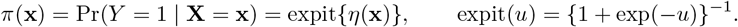

Throughout, logit(*p*) = log{*p*/(1 − *p*)} denotes the log-odds transformation, so that logit{*π*(**x**)} = *η*(**x**) under the data-generating mechanism.

For a given development sample 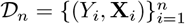 of size *n* generated from the scenario-specific mechanism, we fit the prespecified logistic regression model by maximum likelihood to obtain coefficient estimates 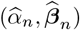. The resulting fitted linear predictor (the *development fit*) evaluated at covariates **x** is

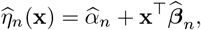

and the corresponding fitted probability is 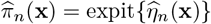. When the dependence on *n* is clear from context, we write 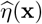 and 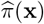 for brevity.

The primary estimand for evaluating overfitting is the logit-scale calibration slope of the development fit when transported to new individuals from the same target population. Let **X**^*∗*^ denote an independent draw from the predictor distribution defining the scenario and let *π*(**X**^*∗*^) be the corresponding true event probability under the data-generating mechanism. We define the population calibration relationship between the true log-odds and the fitted linear predictor as

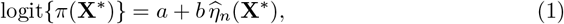

where (*a, b*) are the population least-squares projection coefficients of logit{*π*(**X**^*∗*^)} onto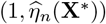. The parameter *b* in (1) is the logit-scale calibration slope estimand used throughout the simulation study. Values *b* < 1 indicate that 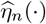 is overly variable relative to the true log-odds, corresponding to overfitting and overly extreme predicted risks; values *b* > 1 indicate insufficient variability in 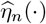 relative to the truth, corresponding to underfitting or excessive shrinkage.

For each scenario and candidate development sample size *n*, we consider repeated independent development samples 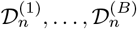 and the associated slopes *b*^(1)^, …, *b*^(*B*)^ implied by (1). The required development sample size, denoted *n*_req_, is defined as the smallest *n* for which the distribution of *b* under repeated sampling satisfies the prespecified calibration-stability criteria in Subsubsection Empirical calibration stability criteria *n*_req_ in Section Methods. The formula-based comparator, denoted *n*_Riley_, is the minimum sample size returned by `pmsampsize` for the same scenario inputs and a fixed global shrinkage target, and is treated as a deterministic function of the design parameters.

### Simulation scenarios

#### Predictor distributions

We considered three data-generating mechanisms (DGMs) for the candidate predictors in order to examine the sensitivity of development sample size recommendations to departures from Gaussian covariate structure. In all scenarios the number of candidate predictors was fixed at *p* = 10, and for each Monte Carlo replicate we generated an independent development sample 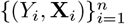 by first simulating predictors **X**_*i*_ = (*X*_*i*1_, …, *X*_*ip*_)^*⊤*^ from the specified DGM and then drawing outcomes from the scenario-specific logistic model described in scenario-specific logistic model described below.

Across all DGMs, predictors were generated as mutually independent within an individual and independent across individuals. Specifically, for each replicate and each *i* = 1, …, *n*, the components *X*_*i*1_, …, *X*_*ip*_ were simulated as independent and identically distributed draws from the DGM-specific marginal distribution. No correlation structure was imposed among predictors (i.e., the working covariance matrix was the identity). This design choice isolates the effect of marginal distributional shape on numerical stability and calibration-related behavior without confounding by multicollinearity. The same independence assumptions were used when generating the large validation covariate set used to evaluate the calibration slope estimand.

Under DGM 1 (Normal), predictors were generated from a standard normal distribution,

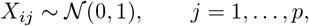

with independence across *j* and across *i*. This mechanism serves as the reference setting in which common analytical approximations for logistic regression performance and information measures are typically most defensible.

Under DGM 2 (Skewed), predictors were generated from an exponential distribution with rate 1,

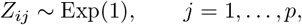

again independently across *j* and *i*. Because the exponential distribution is not centered and has variance 1, we standardized each predictor marginally to ensure comparability of coefficient scaling across DGMs when targeting a common discriminative performance. Specifically, we set

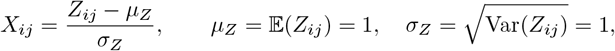

so that *X*_*ij*_ has mean 0 and variance 1 while retaining substantial right-skewness. The same standardization was applied when generating covariates for the calibration step in Subsubsection Calibration of scenario parameters to design targets in Section Methods and for the validation covariate set used in performance evaluation.

Under DGM 3 (Binary), predictors were generated as Bernoulli random variables with success probability 0.5,

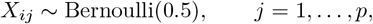

independently across *j* and *i*. This mechanism represents a setting with discrete predictors and limited support, which can amplify separation-like behavior in high-discrimination regimes even when the fitted model is correctly specified.

For all DGMs, the outcome model was fitted in the development sample using the same functional form as the data-generating model (i.e., the fitted logistic regression was correctly specified with respect to the predictors included and their linear effects). Consequently, any discrepancies between formula-based and simulation-based sample size requirements arise from finite-sample behavior, numerical stability, and the adequacy of the approximations underlying analytical criteria, rather than from model misspecification.

#### Signal density

To study whether the distribution of prognostic information across candidate predictors influences development sample size requirements when the nominal model dimension is held fixed, we introduced an explicit parameterization of “signal density.” Let **X** = (*X*_1_, …, *X*_*p*_)^*⊤*^ ∈ ℝ^*p*^ denote the predictor vector for an individual, with *p* = 10 fixed throughout, and let the true linear predictor be

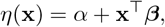

where *α* ∈ ℝ is the intercept and ***β*** ∈ ℝ^*p*^ is the vector of slope coefficients. We parameterize ***β*** as a product of a scalar signal-strength parameter *f* > 0 and a direction vector **d** ∈ ℝ^*p*^:

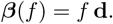

The direction vector **d** determines which predictors carry nonzero effects and their relative magnitudes, while the scalar *f* controls the overall strength of the linear predictor on the logit scale.

We considered two signal-density settings that share the same candidate dimension *p* but differ in how the prognostic signal is allocated across predictors. In the dense-signal setting, all candidate predictors contribute equally to the linear predictor, and we set

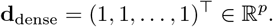

In the sparse-signal setting, only a subset of predictors carries prognostic signal, and we set

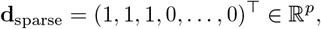

so that exactly three predictors have nonzero coefficients and the remaining seven predictors are pure noise with respect to the outcome. In both cases, the fitted development model includes all *p* predictors, so the number of estimated slope parameters is identical across dense and sparse scenarios and equals *p* (excluding the intercept).

This construction isolates “where the signal lives” while holding nominal model complexity fixed. Because *p* and the fitted model specification are unchanged, any differences in finite-sample behavior between dense and sparse settings arise from the effective allocation of information across parameters rather than from changes in the dimension of the fitted model. Intuitively, under the sparse setting, the true data-generating process has a low-dimensional structure embedded within a higher-dimensional candidate model, whereas under the dense setting, the signal is distributed across all candidate parameters. By subsequently calibrating (*α, f*) within each signal-density setting to achieve common target values of the population prevalence and discrimination (Subsubsection Calibration of scenario parameters to design targets in Section Methods), we ensure that dense and sparse scenarios are comparable with respect to overall outcome frequency and global separability, allowing any residual differences in required development sample size to be attributed specifically to signal density rather than to differences in baseline risk or overall prognostic strength.

#### Calibration of scenario parameters to design targets

For each scenario, outcomes were generated from a correctly specified logistic model with a linear predictor

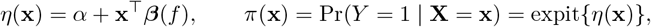

where *α* is the intercept and ***β***(*f*) = *f* **d** is defined by the signal-density specification in Subsubsection Signal density in Section Methods. Scenario parameters were calibrated to satisfy two population-level design targets: the marginal event prevalence *ϕ*_target_ = Pr(*Y* = 1) and the marginal discrimination AUC_target_, defined as the area under the ROC curve for the true risk score *π*(**X**) (equivalently, for the true linear predictor *η*(**X**) under monotone transformation).

Because neither the marginal prevalence nor the AUC has a closed-form expression as a function of (*α, f*) under the non-Gaussian predictor distributions considered, calibration was performed by Monte Carlo integration over a large synthetic covariate population. For each combination of predictor distribution, signal density, and target pair (*ϕ*_target_, AUC_target_), we first generated an independent Monte Carlo sample 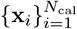 from the scenario-specific covariate distribution, with *N*_cal_ = 1,000,000. This covariate sample was held fixed throughout calibration for that scenario to stabilize the numerical objective functions and ensure deterministic root-finding conditional on {**x**_*i*_}.

Calibration proceeded via nested one-dimensional root finding. For a fixed *f*, the intercept *α* was chosen to match the target prevalence by solving

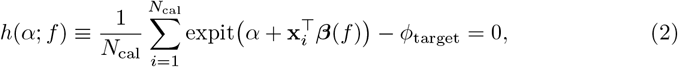

using bisection as implemented in `uniroot`. The root was computed to absolute tolerance 10^*−*8^. Bracketing intervals for *α* were initialized and then adaptively expanded until *h*(*α*_low_; *f*) and *h*(*α*_high_; *f*) had opposite signs, guaranteeing the existence of a solution within the interval under the fixed covariate sample.

Given the resulting prevalence-calibrated intercept *α*(*f*) from (2), the signal-strength parameter *f* was then calibrated to achieve the target discrimination. For any candidate *f*, we computed the deterministic “true” event probabilities 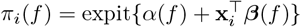 and then evaluated

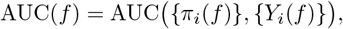

where *Y*_*i*_(*f*) denotes a Bernoulli outcome generated with probability *π*_*i*_(*f*). To eliminate additional Monte Carlo noise from generating *Y*_*i*_(*f*) during calibration, AUC(*f*) was computed using the population risk score *π*_*i*_(*f*) and outcomes drawn once per scenario from Bernoulli(*π*_*i*_(*f*)) using a fixed random seed; sensitivity checks confirmed that AUC(*f*) was stable to this choice given the very large *N*_cal_. The scale parameter *f* was obtained by solving

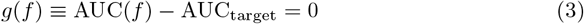

using `uniroot` with absolute tolerance 10^*−*8^. The search interval for *f* was prespecified to cover all target AUC values and expanded if necessary to ensure that *g*(*f*) changed sign over the interval.

Following calibration, we performed validation checks to confirm that the achieved design characteristics matched their targets within the Monte Carlo error. Using an independent covariate sample of size *N*_check_ = 20000 from the same predictor distribution, we recomputed the implied prevalence

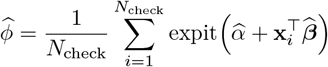

and an empirical AUC based on outcomes generated from Bernoulli 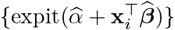. Calibration was accepted if 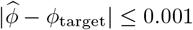 and 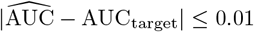 these tolerances were chosen to be substantially smaller than the performance differences of interest in subsequent sample size comparisons. The resulting scenario-specific parameter pairs 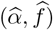 were then used for all downstream simulations. The detailed procedure for this nested root-finding approach is summarized in Algorithm 1.

##### Algorithm 1

Nested Root-Finding Procedure for Parameter Estimation with Monte Carlo Validation

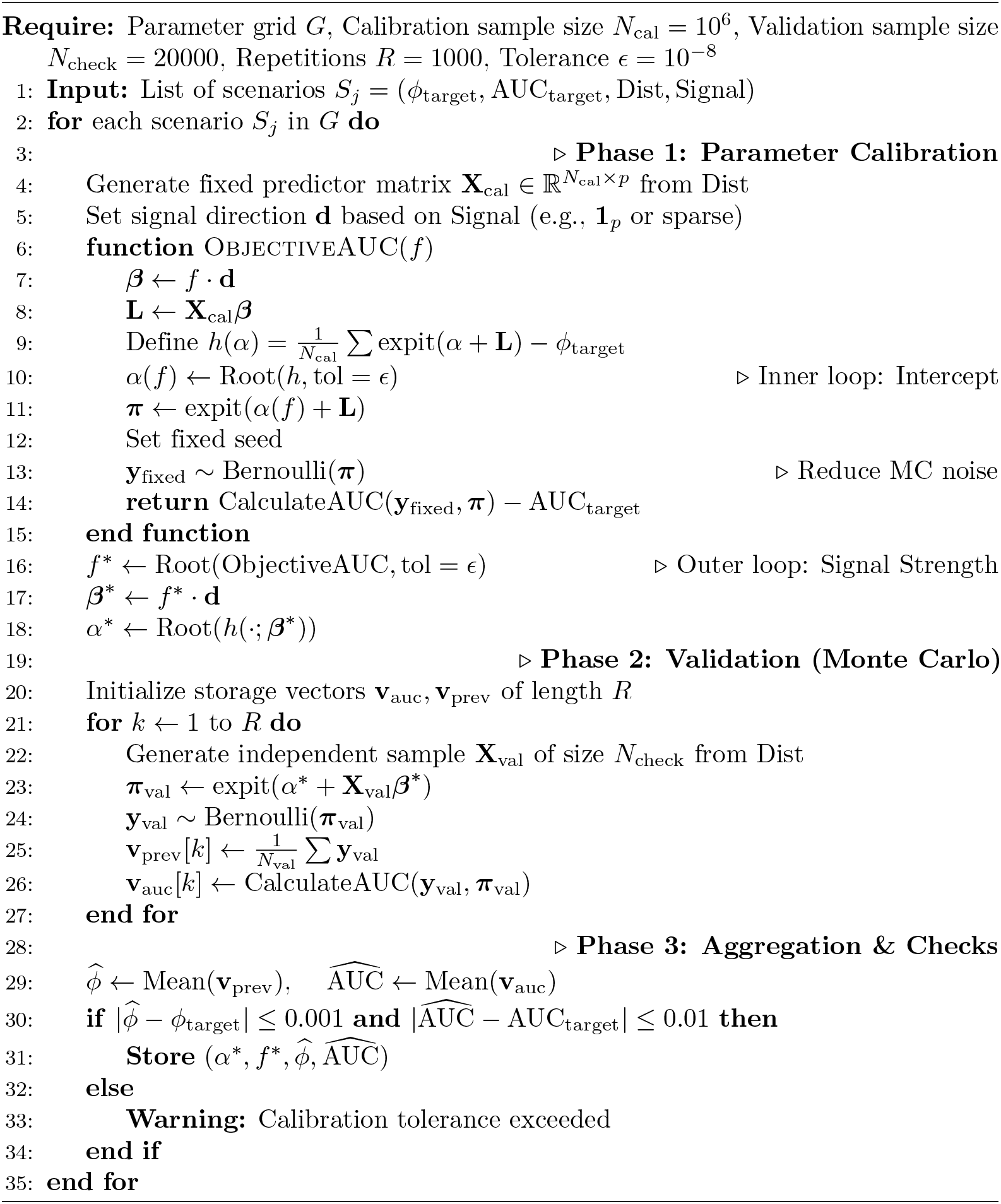

### Evaluation metrics

#### Primary estimand: calibration slope

For a given development dataset 𝒟_*n*_ of size *n*, let 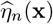 denote the fitted linear predictor from the prespecified logistic regression model, evaluated at covariate vector **x**. Under the scenario-specific data-generating mechanism, the true event probability is *π*(**x**) = Pr(*Y* = 1 | **X** = **x**) with true log-odds logit{*π*(**x**)}. We define the logit-scale population calibration model for predictions from the development fit as

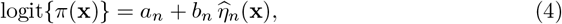

where *a*_*n*_ ∈ ℝ and *b*_*n*_ ∈ ℝ are the population calibration intercept and slope corresponding to the fitted predictor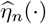. The calibration slope *b*_*n*_ is the primary estimand used throughout this study. Values *b*_*n*_ < 1 indicate that 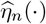 is overly variable relative to the true log-odds, yielding overly extreme predictions on the logit scale (overfitting), whereas *b*_*n*_ > 1 indicates that predictions are insufficiently dispersed relative to the true log-odds (underfitting relative to the data-generating mechanism). The calibration intercept *a*_*n*_ captures miscalibration-in-the-large and equals zero when the average predicted risk agrees with the observed event rate, conditional on the linear predictor scale.

To estimate (*a*_*n*_, *b*_*n*_) with negligible evaluation noise, we use a truth-based procedure that conditions on a large validation covariate sample while avoiding simulation of validation outcomes. Specifically, for each predictor-distribution setting we generate a fixed validation covariate set 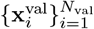 of size *N*_val_ independently from the covariate distribution of the scenario. For each 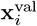, we compute the true probability deterministically as 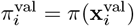 using the known scenario parameters, and form theb true log-odds 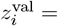 logit 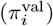. We then estimate (*a*_*n*_, *b*_*n*_) by ordinary least squares regression of 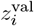 on the fitted linear predictor evaluated on the same covariates, 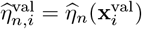:

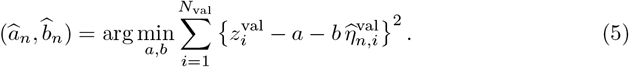

Because 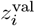 is computed deterministically from the data-generating mechanism, 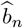 targets the logit-scale calibration slope in (4) without additional Monte Carlo error from simulated validation outcomes. The remaining Monte Carlo variation in 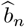 arises solely from the randomness of the development sample 𝒟_*n*_ (through 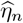) and, to a negligible degree, from the finite but large *N*_val_.

This truth-based logit-scale slope is directly aligned with the conceptual target of controlling overfitting in the fitted linear predictor. In particular, Riley-type minimum sample size criteria enforce a prespecified global shrinkage factor *S* for maximum-likelihood logistic regression fits. Under correct model specification and standard regularity conditions, the global shrinkage factor can be interpreted as an approximation to the expected out-of-sample logit-scale calibration slope of the fitted linear predictor. Accordingly, comparing the empirical behavior of *b*_*n*_ to a target such as *S* = 0.90 is meaningful at the level of expected performance, while recognizing that numerical instability and violations of regularity in high-discrimination regimes may weaken this approximation; for that reason, our primary evaluation is based on the directly estimated *b*_*n*_ defined in (4).

### Separation and convergence diagnostics

To investigate whether numerical pathologies in maximum-likelihood logistic regression contribute to the observed discrepancy between *n*_Riley_ and *n*_req_ in high-discrimination settings, we quantified separation- and convergence-related diagnostics in the development fits. The purpose of these diagnostics was mechanistic rather than inferential: they were used to document how often standard likelihood-based regularity conditions appear to be violated at the formula-recommended sample size, which may undermine analytical approximations that rely on well-behaved maximum-likelihood estimates.

Diagnostics were evaluated for unpenalized logistic regression fitted by maximum likelihood at *n* = *n*_Riley_ for each scenario. For replicate **r**, let 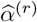 and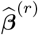 denote the fitted intercept and slope vector, and let 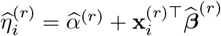 be the fitted linear predictor for individual *i* = 1, …, *n* in the development sample. The corresponding fitted risks are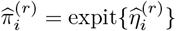. We recorded the following replicate-level indicators and summarized them as proportions across Monte Carlo replicates.

Non-convergence was defined as the failure of the fitting routine to declare convergence within the maximum number of iterations and/or production of non-finite coefficient estimates. Operationally, the indicator was set to one if the model fit returned a non-zero convergence code, if any element of 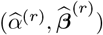 the fitting call produced an error that prevented extraction of coefficients. This definition is intentionally conservative, as non-finite estimates and optimizer failure both indicate that the likelihood surface is not being reliably optimized under the chosen numerical routine.

A separation warning indicator was constructed to capture quasi-complete or complete separation manifested as extreme fitted log-odds or fitted probabilities, even when the optimizer reports convergence. For replicate *r*, we defined

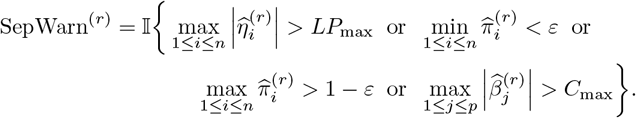

We set the thresholds to *LP*_max_ = 12, *ε* = 10^*−*6^, and *C*_max_ = 5, with the coefficient threshold applied only to slope coefficients (excluding the intercept). The log-odds threshold corresponds to fitted probabilities of approximately expit(±12) ≈ (6 × 10^*−*6^, 0.999994), and thus identifies near-deterministic predictions that are typically regarded as numerically extreme in logistic regression computations. The probability threshold *ε* = 10^*−*6^ is aligned with common numerical conventions for defining fitted probabilities effectively equal to 0 or 1. The coefficient-magnitude threshold *C*_max_ = 5 was chosen as a pragmatic marker of large effect sizes on the log-odds scale that, in the context of standardized predictors, often accompany separation-driven instability; it is not intended as a substantive clinical threshold, but as a numerical diagnostic that is comparable across scenarios.

Because the composite separation-warning indicator can be triggered by different phenomena, we additionally reported a coefficient-divergence indicator to isolate the contribution of large estimated slopes:

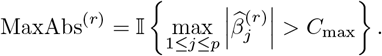

This was summarized as the proportion of replicates in which at least one slope coefficient exceeded *C*_max_ in absolute value.

We also recorded an extreme fitted-risk event indicator, defined at the replicate level as

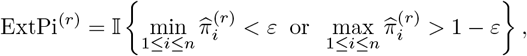

and reported the corresponding empirical probability across replicates. This quantity differs from the separation-warning indicator in that it focuses specifically on the presence of near-deterministic fitted risks in the development sample, which is the most direct operational manifestation of separation in terms of predictions.

Finally, we defined a non-estimable calibration slope indicator to capture rare cases in which the truth-based logit-scale calibration slope could not be computed as a finite number. For replicate *r*, the indicator NonEstSlope^(*r*)^ was set to one if the calibration-slope estimation procedure returned a non-finite value (e.g., due to missing coefficients from the development fit, numerical overflow, or a degenerate fitted linear predictor). When this occurred, the replicate was treated conservatively as a failure for the tail-risk criterion used to define *n*_req_.

These thresholds were pre-specified and held fixed across all scenarios to ensure comparability of diagnostic rates. They are intended to flag numerically extreme behavior that indicates potential violations of standard regularity conditions underlying likelihood-based approximations, rather than to provide a formal classification of separation in the strict theoretical sense. The resulting diagnostic frequencies at *n* = *n*_Riley_ were then used to interpret whether high-discrimination discrepancies coincide with an increased prevalence of separation-like numerical behavior.

### Determination of sample sizes

#### Formula-based benchmark (*n*_Riley_)

For each simulation scenario, we computed a formula-based minimum development sample size using the approach proposed by Riley and colleagues for prediction models with a binary outcome and fitted by maximum-likelihood logistic regression (Riley et al., 2019). We denote this benchmark by *n*_Riley_ and obtain it using the `pmsampsize` R package (Ensor et al., 2023), which implements the closed-form criteria of Riley et al. (2019) for minimum sample size planning.

The required inputs to `pmsampsize` were specified to match the scenario design targets. Let *p* denote the total number of candidate predictor parameters to be estimated in the development model, excluding the intercept. In the simulation study, *p* = 10 for all scenarios. Let *ϕ*_target_ denote the target event prevalence and let AUC_target_ denote the target C-statistic (area under the ROC curve) that characterizes the anticipated discrimination of the developed model in the target population. For each combination of (*ϕ*_target_, AUC_target_), `pmsampsize` internally converts the anticipated discrimination into an anticipated Cox–Snell *R*^2^ measure, 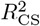, using its built-in mapping between the C-statistic and explained variation for logistic regression. On the basis of (*p, ϕ*_target_, 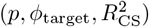) and a user-specified shrinkage target, the routine returns the recommended minimum development sample size *n*_Riley_ as the maximum of the sample sizes required to satisfy the three Riley criteria: (i) a lower bound on the global shrinkage factor, (ii) a limit on optimism in model fit, and (iii) adequate precision for estimation of the intercept (calibration-in-the-large).

In this study, we set the global shrinkage target to *S* ≥ 0.90 for all scenarios, consistent with the common recommendation that the average calibration slope (or equivalently the average shrinkage of the linear predictor under regularity) should be close to one and not materially below 0.90. The resulting value *n*_Riley_ therefore represents the minimum sample size recommended by the Riley framework, as operationalized in `pmsampsize`, for an unpenalized logistic regression model with *p* candidate parameters, anticipated discrimination AUC_target_, and outcome prevalence *ϕ*_target_.

A key structural property of this benchmark is that it depends on the number of candidate parameters *p* and the anticipated overall model performance (via 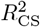, itself derived from AUC_target_ and *ϕ*_target_), but it does not condition on how prognostic signal is distributed across predictors. In our simulation design, dense and sparse signal structures share the same *p* by construction, and scenario calibration ensures the same (*ϕ*_target_, AUC_target_) (hence the same anticipated 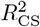) across signal-density conditions within a given predictor-distribution setting. Consequently, for fixed (*p, ϕ*_target_, AUC_target_), `pmsampsize` yields the same *n*_Riley_ for dense and sparse scenarios, and any observed differences in empirical required sample size across signal-density conditions arise from finite-sample behavior rather than from differences in the formula inputs.

#### Empirical calibration stability criteria *n*_req_

For each scenario in the factorial design, we define the required development sample size, denoted *n*_req_, as the smallest integer *n* for which a prespecified logit-scale calibration-stability criterion is achieved under repeated sampling of development datasets from the data-generating mechanism.

Let 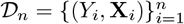 denote a randomly drawn development sample of size *n* from the scenario-specific joint distribution of (*Y*, **X**), and let 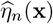 denote the fitted linear predictor obtained by fitting the prespecified (correctly specified) logistic regression model to 𝒟_*n*_ using maximum likelihood. Let *b*_*n*_ denote the corresponding out-of-sample calibration slope for predictions from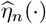, defined as the slope parameter in the population calibration relationship

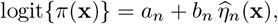

where *π*(**x**) = Pr(*Y* = 1 | **X** = **x**) is the true event probability under the data-generating mechanism and *a*_*n*_ is an intercept term. Here *b*_*n*_ is a functional of the random development sample 𝒟_*n*_ through its dependence on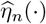.

All probability statements and expectations below are taken with respect to the randomness in repeated draws of the development dataset 𝒟_*n*_ from the scenario-specific data-generating mechanism (i.e., repeated model development). In particular, 𝔼 (*b*_*n*_) denotes the mean calibration slope across repeated development samples of size n, and Pr(*b*_*n*_ < 0.80) denotes the probability that a model developed on a random sample of size *n* yields a calibration slope below 0.80 when evaluated out of sample under the truth-based calibration framework.

We define *n*_req_ as the smallest *n* such that both of the following conditions hold:

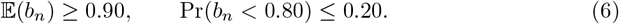

The first condition targets control of average overfitting on the logit scale, while the second condition constrains the tail risk of severe miscalibration due to instability in the fitted linear predictor. In the simulation study, both quantities were estimated empirically via Monte Carlo replication at candidate sample sizes *n*, and *n*_req_ was obtained as the minimum n satisfying (6) within Monte Carlo uncertainty within Monte Carlo uncertainty (described below).

#### Search algorithm for *n*_req_

For each scenario defined by the predictor distribution, signal-density structure, target discrimination, and target prevalence, we estimated the required development sample size *n*_req_ by a simulation-based bisection algorithm. The bisection approach was adopted to avoid an exhaustive grid search over candidate sample sizes while still identifying the minimum *n* that satisfies the calibration-stability criteria in criteria in Equation 6. All probability statements in this section are with respect to repeated sampling of the development dataset under the scenario-specific data-generating mechanism.

For a given scenario, we defined an initial search interval [*n*_min_, *n*_max_] with *n*_min_ = 100 and *n*_max_ = 5000. At each bisection step, we set *n*_mid_ = ⌊(*n*_min_ + *n*_max_)/2⌋ and evaluated whether *n*_mid_ met the prespecified criteria. If *n*_mid_ was deemed acceptable, we updated the upper bound to *n*_max_ ← *n*_mid_; otherwise, we updated the lower bound to *n*_min_ ← *n*_mid_. The procedure terminated when the interval width satisfied *n*_max_ − *n*_min_ ≤ 25, at which point we reported *n*_req_ = *n*_max_ as the smallest sample size supported by the evidence accumulated under the decision rule described below.

Evaluating a candidate *n* required repeated Monte Carlo replicates. For replicate *r* = 1, …, B, we generated an independent development dataset of size *n*, fitted the prespecified unpenalized logistic regression model by maximum likelihood, and obtained the fitted linear predictor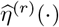. Calibration was then evaluated using the truth-based logit-scale calibration slope defined in Subsubsection Primary estimand: calibration slope in Section Methods, computed on a large, fixed validation covariate set of size *N*_val_ = 50,000 generated once per predictor-distribution setting and reused across scenarios to remove incidental variation due to differing validation case-mix. This produced replicate-specific slope estimates *b*^(*r*)^, from which we computed the empirical mean 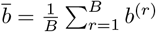 and the empirical failure proportion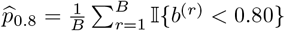, where non-estimable slopes (if any) were conservatively classified as failures.

##### Algorithm 2

Simulation-based Bisection Search for Required Sample Size (*n*_req_)

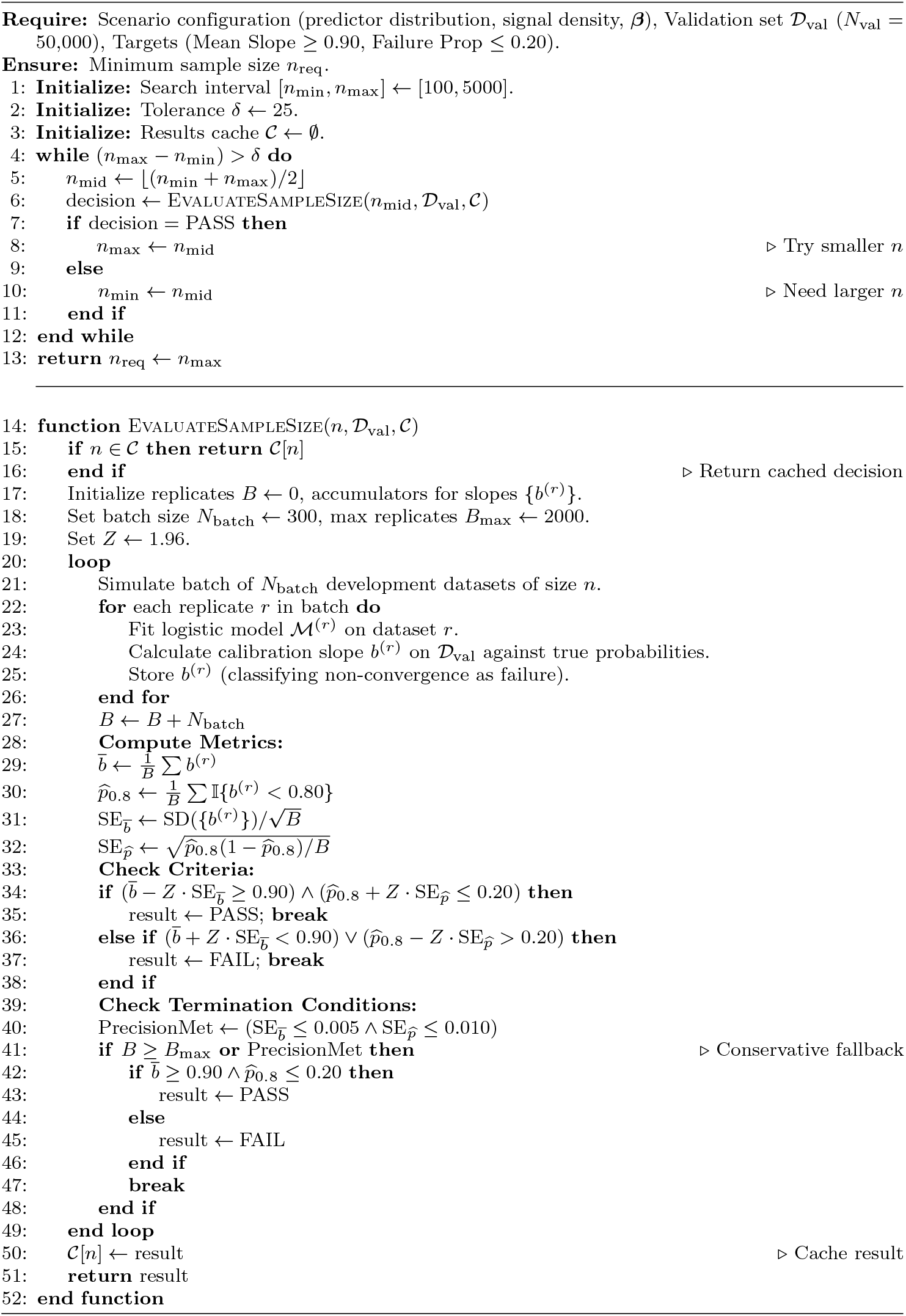

To control Monte Carlo uncertainty, each evaluation began with *B* = 300 replicates and was extended adaptively in batches of 300 up to a maximum of *B*_max_ = 2000 replicates when needed. After each batch, we computed Monte Carlo standard errors for both criteria. For the mean slope, we used

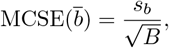

where *s*_*b*_ is the sample standard deviation of 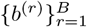 . For the failure proportion, we used the binomial Monte Carlo standard error

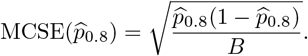

We then formed conservative uncertainty bounds using a normal-approximation multiplier of 1.96 to summarize the level of Monte Carlo uncertainty without invoking hypothesis testing. A candidate n was classified as acceptable if both of the following held:

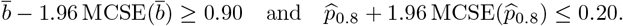

Conversely, a candidate n was classified as unacceptable if either of the following held:

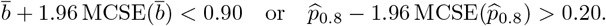

If neither classification was supported because the bounds overlapped the target region, additional batches were simulated until classification was possible, the replicate maximum *B*_max_ was reached, or pre-specified Monte Carlo precision thresholds were met. Precision thresholds were set to 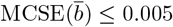 and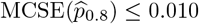. If these thresholds were met before a definitive classification, we applied a conservative rule by requiring the point estimates to satisfy both targets, i.e., 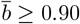 and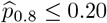, to avoid declaring adequacy on the basis of imprecise Monte Carlo evidence.

To improve computational efficiency, we cached the full set of replicate results for each evaluated (scenario, *n*) combination so that repeated visits to the same *n* during bisection did not trigger re-simulation. All simulation was executed in parallel across replicates within each batch. Reproducibility under parallel execution was ensured using the `L’Ecuyer--CMRG` random number generator with deterministic seeding based on the scenario identifier, candidate *n*, and batch index, yielding reproducible replicate streams regardless of the number of worker processes or their scheduling. The combination of bisection search, adaptive replication, and caching provided a computationally tractable approach for estimating *n*_req_ over the full factorial design. The complete logic for this estimation process, including the specific decision boundaries and adaptive stopping rules, is formalized in Algorithm 2.

## Results

### Validation of calibration step (targets achieved)

Before evaluating sample size requirements, we verified that the scenario-calibration procedure described in Subsubsection Calibration of scenario parameters to design targets in Section Methods produced data-generating mechanisms that matched the prespecified design targets for both outcome prevalence and discrimination. For each combination of predictor distribution, signal density, target prevalence *ϕ*_target_ ∈ {0.05, 0.10, 0.20}, and target discrimination AUC_target_ ∈ {0.70, 0.75, 0.80, 0.85, 0.90}, we calibrated (α, f) using a large Monte Carlo covariate sample and then evaluated the achieved population characteristics on an independent Monte Carlo sample to avoid reusing calibration noise.

Across the full factorial grid, the achieved event prevalence closely matched the target value. The absolute deviation between the Monte Carlo estimate of 𝔼{*π*(**X**)} and *ϕ*_target_ was typically on the order of 10^*−*3^ or smaller, with Monte Carlo standard errors that were negligible relative to this tolerance. Likewise, the achieved discrimination closely matched the target AUC. The Monte Carlo AUC estimates were within approximately 10^*−*2^ of AUC_target_ throughout, consistent with the expected finite Monte Carlo error from estimating a rank-based functional under a fixed covariate-generating distribution. These checks confirm that the calibration algorithm reliably produced scenarios with the intended prevalence and discriminative strength, so that subsequent discrepancies between *n*_req_ and *n*_Riley_ can be attributed to the sampling behavior of the development estimator under the specified design conditions rather than to miscalibration of the underlying data-generating parameters.

Table 1 summarizes the validation results by reporting, for each scenario, the target prevalence and AUC alongside their Monte Carlo estimates (with corresponding standard errors), together with the calibrated intercept *α* and signal-strength factor *f* that define the scenario-specific outcome model. The tight agreement between targets and Monte Carlo estimates demonstrates that the parameter calibration step achieved its purpose and provides an empirically verified foundation for the sample size comparisons reported in the remainder of the Results section.

**Table 1.**
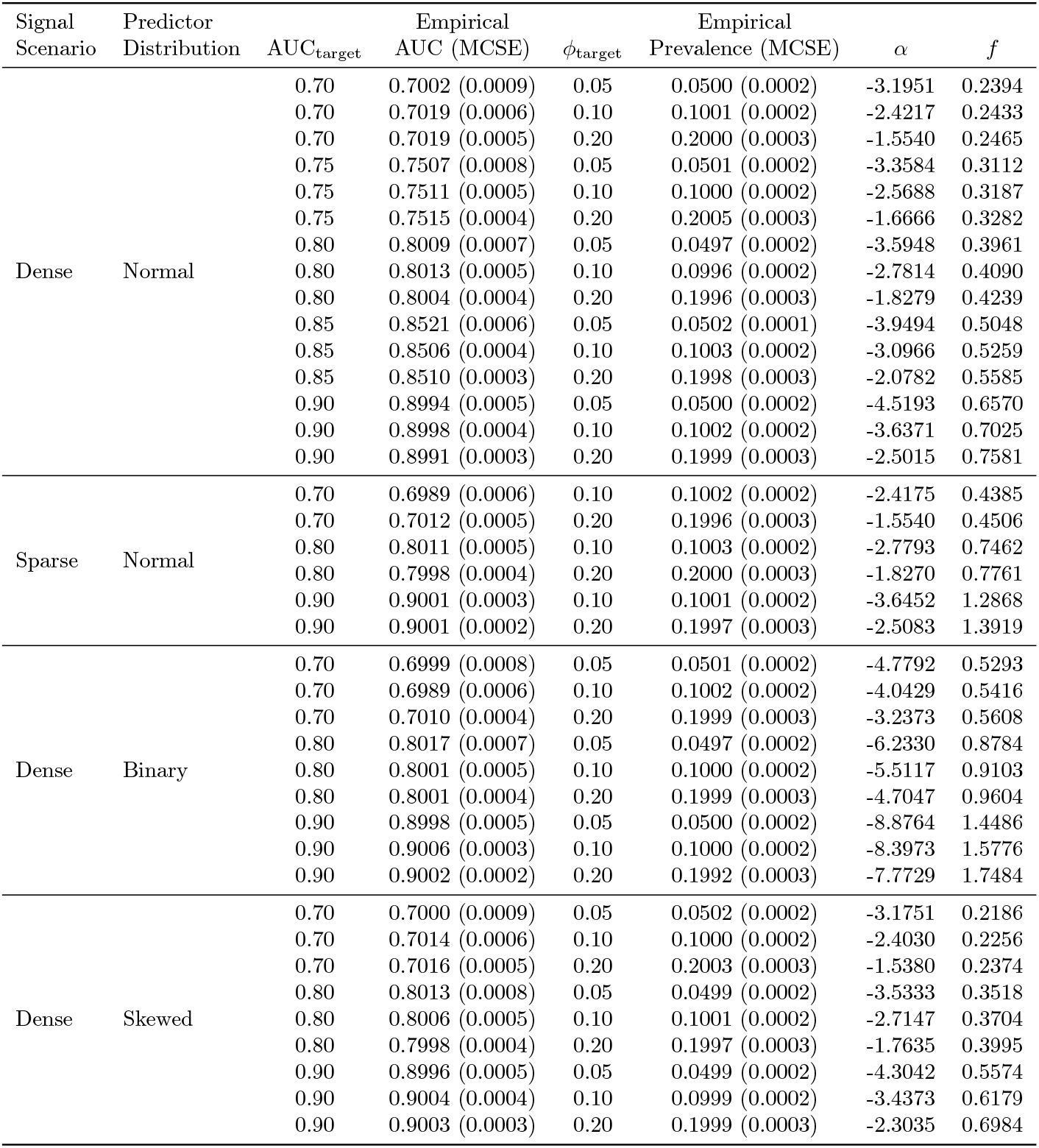
Validation of calibrated scenario targets. For each scenario, the target AUC and prevalence are reported alongside Monte Carlo estimates (Monte Carlo standard errors in parentheses), together with the calibrated intercept α and signal-strength factor f.

### Primary result: discrepancy as a function of discrimination (AUC)

The primary comparison of interest was between the simulation-based required development sample size *n*_req_ and the formula-based benchmark *n*_Riley_, evaluated as a function of target discrimination AUC_target_. For each scenario, *n*_Riley_ was obtained from `pmsampsize` under the prespecified shrinkage target *S* ≥ 0.90, and *n*_req_ was defined as the smallest *n* satisfying the calibration-stability criteria 𝔼 (*b*) ≥ 0.90 and Pr(*b* < 0.80) ≤ 0.20 under repeated development sampling (Subsubsection Empirical calibration stability criteria *n*_req_ in Section Methods). Fig 2 and Table 2 summarize the resulting relationship between discrimination and the gap *n*_req_ − *n*_Riley_.

**Table 2.** Comparison of simulation-based required sample sizes (*n*_req_) versus the Riley benchmark (*n*_Riley_) across target prevalence and discrimination in the Normal-predictor, Dense-signal scenarios. Relative difference is (*n*_Riley_ − *n*_req_)/*n*_req_.

| $\phi_{\text{target}}$ | $\text{AUC}_{\text{target}}$ | $n_{\text{req}}$ | $n_{\text{Riley}}$ | $n_{\text{req}} - n_{\text{Riley}}$ | Rel. Diff. |
| --- | --- | --- | --- | --- | --- |
| 0.05 | 0.70 | 3124 | 3494 | -370 | 11.8% |
|  | 0.75 | 2091 | 2129 | -38 | 1.8% |
|  | 0.80 | 1574 | 1395 | 179 | -11.4% |
|  | 0.85 | 1248 | 946 | 302 | -24.2% |
|  | 0.90 | 1095 | 652 | 443 | -40.5% |
| 0.10 | 0.70 | 1746 | 1857 | -111 | 6.4% |
|  | 0.75 | 1267 | 1143 | 124 | -9.8% |
|  | 0.80 | 904 | 755 | 149 | -16.5% |
|  | 0.85 | 769 | 519 | 250 | -32.5% |
|  | 0.90 | 693 | 366 | 327 | -47.2% |
| 0.20 | 0.70 | 1037 | 1051 | -14 | 1.4% |
|  | 0.75 | 750 | 655 | 95 | -12.7% |
|  | 0.80 | 559 | 437 | 122 | -21.8% |
|  | 0.85 | 501 | 305 | 196 | -39.1% |
|  | 0.90 | 482 | 255 | 227 | -47.1% |

**Fig 2.**
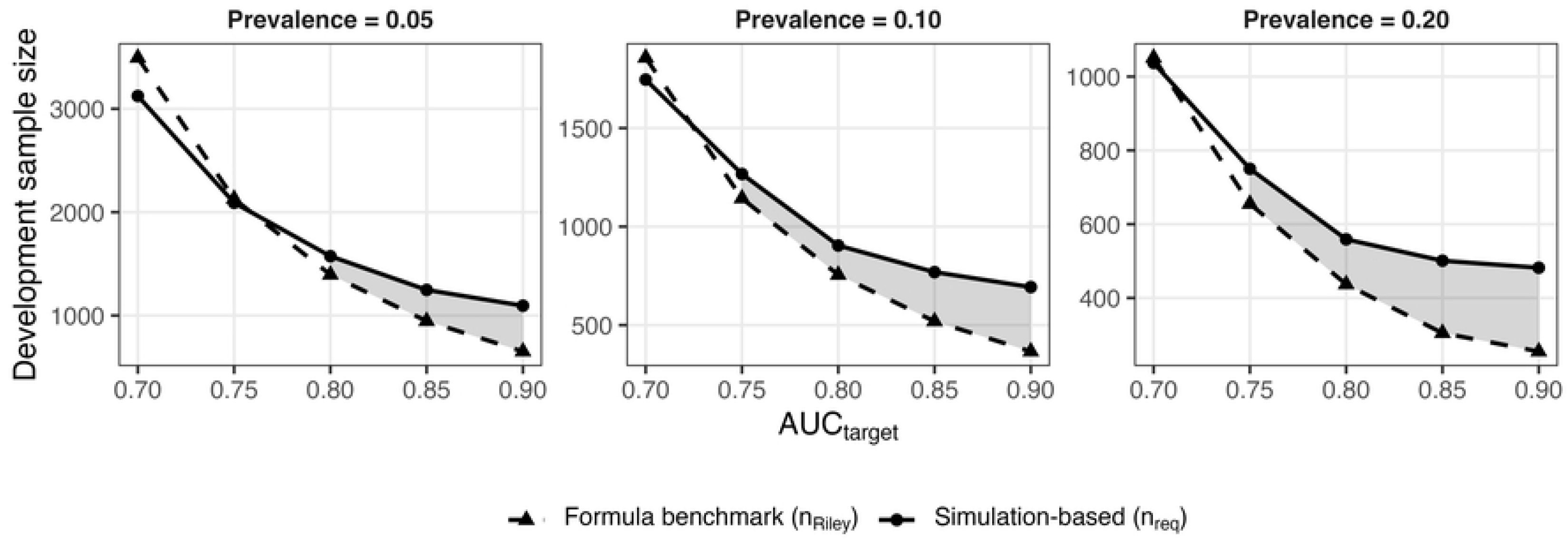
Sample Size Discrepancy: Simulation vs. Formula. The plot illustrates the required sample size for a model with p=10 normally distributed predictors. Shaded areas represent the deficit in sample size when using the analytical formula compared to the simulation-based requirement.

At moderate discrimination, the closed-form benchmark provided a close approximation to the simulation-based requirement. In particular, at AUC_target_ = 0.75 the two approaches yielded similar required sample sizes, with relative differences that were small in magnitude. This indicates that, in settings with moderate discrimination, the combination of the shrinkage-based requirement and the ancillary criteria embedded in `pmsampsize` leads to sample size recommendations that align well with directly evaluated logit-scale calibration stability under repeated development sampling.

As discrimination increased, a systematic discrepancy emerged. From AUC_target_ = 0.80 onward, *n*_Riley_ increasingly underestimated *n*_req_, and the magnitude of underestimation grew monotonically with AUC_target_. The pattern was most pronounced at the upper end of the discrimination range. For example, in the normal-predictor, dense-signal scenarios, relative differences progressed from modest underestimation at AUC_target_ = 0.80 to substantial underestimation at AUC_target_ = 0.90 (Table 2). These results demonstrate that, although both approaches imply that fewer observations are required as discrimination increases, the formula-based benchmark decreases more rapidly than the empirically required sample size for controlling calibration slope behavior under repeated development sampling.

Fig 3 illustrates the practical consequences of these sample size discrepancies on model calibration. By design, the simulation-derived sample sizes (*n*_req_, pink boxplots) maintained a mean calibration slope of approximately 0.90 across all levels of discrimination, satisfying the stability definition. In contrast, models developed using the benchmark sample sizes (*n*_Riley_, blue boxplots) exhibited a progressive deterioration in calibration performance as AUC_target_ increased. While performance was comparable at AUC_target_ = 0.75, the distributions for *n*_Riley_ shifted systematically downward at higher discrimination levels. At AUC_target_ = 0.90, the average calibration slope for the benchmark group dropped substantially below the 0.90 threshold (e.g., mean slope ≈ 0.80 for *ϕ*_target_ = 0.20), indicating that the formula-based sample sizes were insufficient to counteract the overfitting induced by high signal separation.

**Fig 3.**
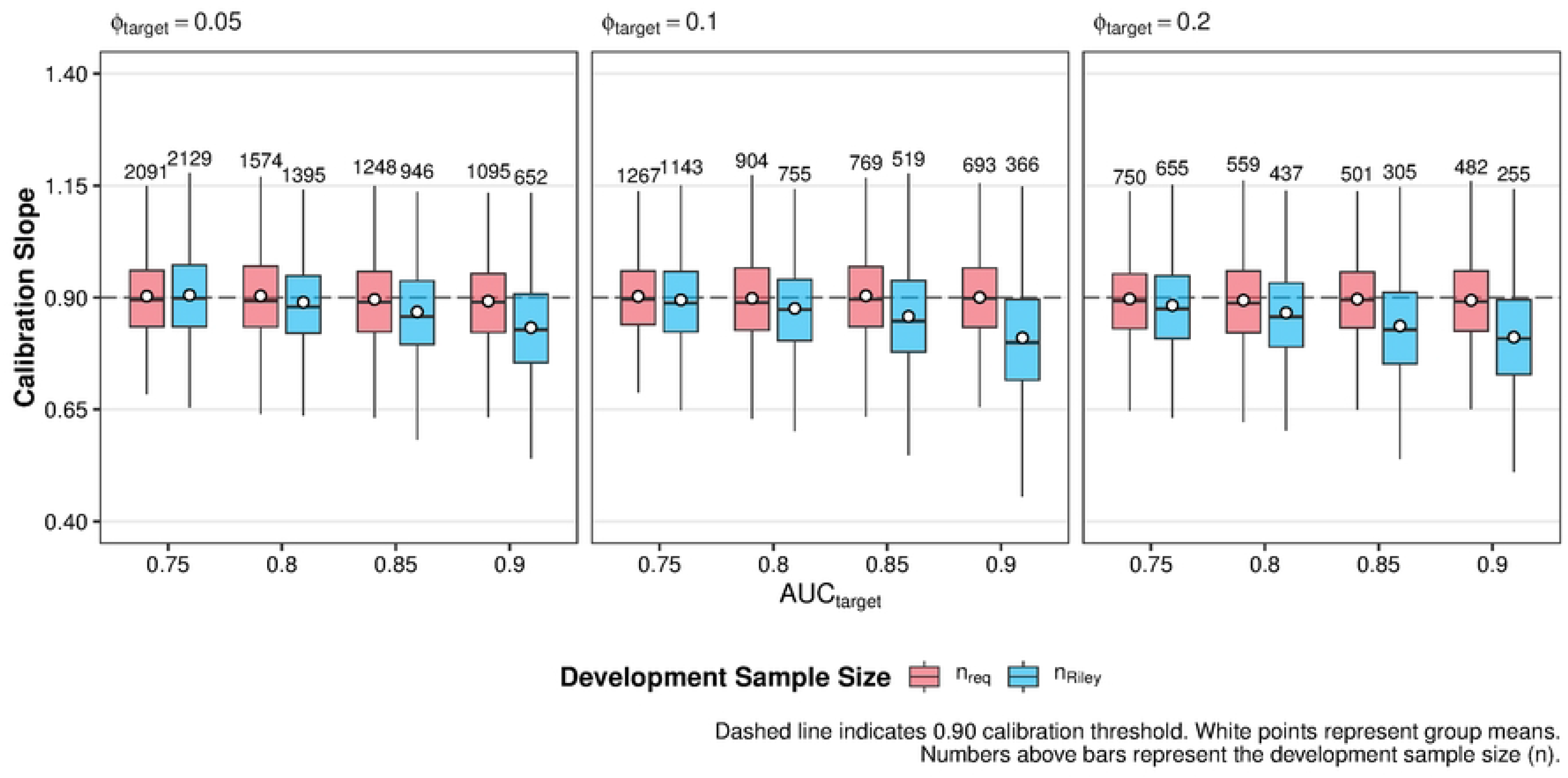
Distribution of validation calibration slopes for logistic regression models developed using simulation-based (*n*_req_) versus formula-based (*n*_Riley_) sample sizes. Results are stratified by target prevalence (*ϕ*_target_ ∈ {0.05, 0.10, 0.20}) and target discrimination (AUC_target_ ∈ {0.75, 0.80, 0.85, 0.90} with p=10 normally distributed predictors). For each scenario, models were developed using sample sizes defined by the simulation requirement (*n*_req_, pink) or the Riley method (*n*_Riley_, blue) using the underlying data-generating parameters (*α, f*) detailed in Table 1. The boxplots display the variability of calibration slopes estimated in independent validation data across simulation repetitions. The dashed horizontal line marks the shrinkage target of 0.90; white points represent group means. Numbers displayed above the whiskers indicate the specific development sample size (*n*) used for that condition.

Overall, the primary result is a discrimination-dependent divergence: agreement between *n*_req_ and *n*_Riley_ is acceptable at moderate AUC_target_, whereas the benchmark shows a consistent and increasing tendency to recommend smaller sample sizes than are required to meet the prespecified calibration-stability targets as AUC_target_ becomes high. This discrepancy motivates the subsequent mechanism analyses, which examine whether numerical and information-based regularity conditions assumed by the shrinkage approximation remain adequate as discrimination increases.

### Mechanism results: separation-like irregularity at *n* = *n*Riley

To assess whether the discrimination-dependent discrepancy between *n*_req_ and *n*_Riley_ could be explained by violations of numerical regularity at the formula-recommended sample size, we evaluated separation and convergence diagnostics for unpenalized logistic regression fits at *n* = *n*_Riley_ (Subsubsection Separation and convergence diagnostics in Section Methods). Table 3 reports the empirical frequencies of non-convergence, separation warnings, and extreme fitted-risk behavior across Monte Carlo replicates for the normal-predictor, dense-signal scenarios, indexed by *ϕ*_target_ and AUC_target_.

**Table 3.** Separation and convergence diagnostics for unpenalized logistic regression fits at *n* = *n*_Riley_ by target prevalence and discrimination. Values are proportions across Monte Carlo replicates. “Separation warning” denotes a replicate-level quasi-separation indicator triggered if max_*i*_ 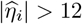, or if any fitted probability satisfies 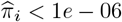 or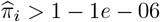, or if max_*j*_ 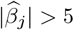 (excluding the intercept). “Extreme π rate” is the probability that at least one individual in the replicate has 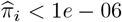 or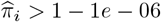 .

| $\phi_{\text{target}}$ | $\text{AUC}_{\text{target}}$ | Non-converged | Separation warning | $\max_j \vert \hat{\beta}_{n,j}\vert > C_{\text{max}}$ | Extreme $\hat{\pi}$ rate | Non-estimable slope |
| --- | --- | --- | --- | --- | --- | --- |
| 0.05 | 0.70 | 0 | 0.000 | 0 | 0.000 | 0 |
|  | 0.75 | 0 | 0.000 | 0 | 0.000 | 0 |
|  | 0.80 | 0 | 0.001 | 0 | 0.000 | 0 |
|  | 0.85 | 0 | 0.045 | 0 | 0.004 | 0 |
|  | 0.90 | 0 | 0.497 | 0 | 0.191 | 0 |
| 0.10 | 0.70 | 0 | 0.000 | 0 | 0.000 | 0 |
|  | 0.75 | 0 | 0.000 | 0 | 0.000 | 0 |
|  | 0.80 | 0 | 0.000 | 0 | 0.000 | 0 |
|  | 0.85 | 0 | 0.018 | 0 | 0.002 | 0 |
|  | 0.90 | 0 | 0.349 | 0 | 0.128 | 0 |
| 0.20 | 0.70 | 0 | 0.000 | 0 | 0.000 | 0 |
|  | 0.75 | 0 | 0.000 | 0 | 0.000 | 0 |
|  | 0.80 | 0 | 0.000 | 0 | 0.000 | 0 |
|  | 0.85 | 0 | 0.005 | 0 | 0.002 | 0 |
|  | 0.90 | 0 | 0.197 | 0 | 0.068 | 0 |

Across all discrimination levels considered, outright optimizer failure was not observed at *n* = *n*_Riley_ (Non-converged = 0 throughout Table 3). Moreover, the coefficient-magnitude flag max_*j*_ 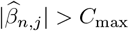 did not trigger under the prespecified threshold *C*_max_ = 5 (excluding the intercept), indicating that the observed irregularities were not primarily expressed as extreme coefficient divergence at this particular cutoff. In contrast, the replicate-level separation warning indicator, defined as

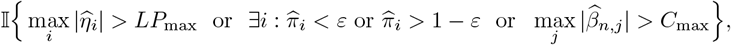

with *LP*_max_ = 12 and *ε* = 10^*−*6^, increased sharply with discrimination. Separation warnings were essentially absent at AUC_target_ ≤ 0.80 but became non-negligible by AUC_target_ = 0.85 and common at AUC_target_ = 0.90, where the warning frequency reached 0.524 when *ϕ*_target_ = 0.05, 0.346 when *ϕ*_target_ = 0.10, and 0.197 when *ϕ*_target_ = 0.20 (Table 3). This pattern demonstrates that, even when standard fitting routines report nominal convergence, the fitted linear predictor can enter regimes indicative of quasi-separation as discrimination increases.

Consistent with this interpretation, extreme fitted risks became increasingly frequent at high discrimination. The replicate-level extreme 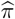 rate, defined as the probability that at least one individual in the development sample attains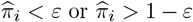, rose markedly with AUC_target_. At AUC_target_ = 0.90, the extreme 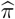 rate was 0.208 for *ϕ*_target_ = 0.05, 0.142 for *ϕ*_target_ = 0.10, and 0.067 for *ϕ*_target_ = 0.20, whereas it was near zero at lower discrimination levels (Table 3). Importantly, the truth-based logit-scale calibration slope remained numerically estimable in all replicates under the present settings (Non-estimable slope = 0 throughout), implying that the observed instability manifested as extreme predicted risks and separation-like behavior rather than complete failure of the calibration-slope estimand.

Taken together, Table 3 indicates that separation-like irregularity at *n* = *n*_Riley_ is strongly discrimination-dependent: non-convergence is rare, but separation warnings and extreme risk predictions become increasingly common as AUC_target_ increases. This finding is directly relevant to the interpretation of the closed-form shrinkage approximation underpinning *n*_Riley_. Riley-type criteria use likelihood-based information measures (e.g., Δχ^2^ and derived shrinkage approximations) that are justified under standard regularity conditions for maximum-likelihood estimation, including the existence and stability of the MLE and well-behaved curvature of the log-likelihood. The observed increase in separation-like behavior at high discrimination provides empirical evidence that these regularity conditions are frequently strained at *n* = *n*_Riley_, even in the absence of explicit non-convergence, offering a plausible mechanism for why the formula-based benchmark becomes optimistic as discrimination increases.

### Secondary results

Across the full factorial grid, we examined whether departures from multivariate normal predictors and variation in signal density materially altered the discrimination-driven discrepancy reported above. Overall, these secondary analyses indicated that the primary qualitative pattern was robust: for fixed candidate parameter dimension, the relationship between discrimination and the gap *n*_req_ − *n*_Riley_ was not materially altered by either predictor distribution or signal-density specification.

First, sensitivity to predictor distribution was modest (Table 4). Under the dense-signal scenarios, simulation-derived required sample sizes *n*_req_ were similar when predictors were generated as standard normal, standardized skewed continuous variables, or Bernoulli(0.5) indicators, after calibrating (*α, f*) to the same target prevalence and discrimination. Differences in *n*_req_ across distributions were most apparent at lower discrimination, where skewed predictors occasionally yielded slightly smaller *n*_req_ than normal or binary predictors, but these deviations were not systematic and narrowed at higher discrimination. In all distributions, the achieved mean calibration slopes at *n*_req_ remained close to the design target and the reliability constraint Pr(*b* < 0.80) ≤ 0.20 was satisfied, indicating that the bisection procedure delivered comparable calibration stability across data-generating mechanisms.

**Table 4.**
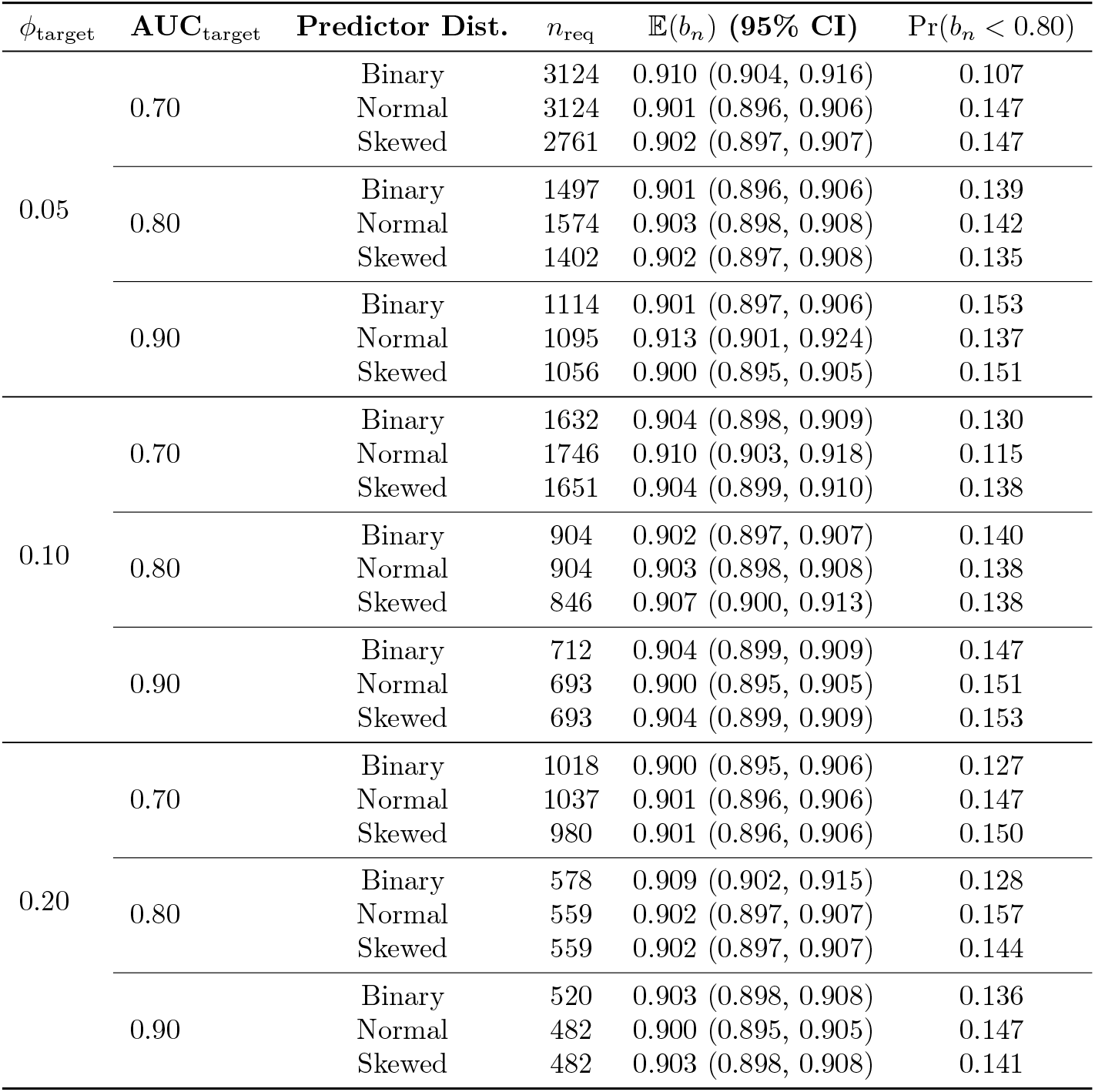
Simulation-based required sample size (dense-signal scenario) by predictor distribution, prevalence, and target AUC. Targets were 𝔼 (*b*_*n*_) ≥ 0.90 and Pr(*b*_*n*_ < 0.80) ≤ 0.20.

Second, sensitivity to signal density was limited when the total number of candidate parameters was held fixed (Table 5. Comparing dense (signal distributed across all *p* predictors) versus sparse (signal concentrated in a subset) configurations, the simulation-based *n*_req_ values differed only slightly and without a consistent direction across settings. In particular, for the prevalences considered and *p* = 10, changing where signal resided affected neither the qualitative discrimination trend nor the overall magnitude of n_req_ beyond small fluctuations. This empirical finding is consistent with the fact that the formula benchmark *n*_Riley_ depends on *p*, the anticipated overall model fit, and the prevalence, but does not condition on how prognostic information is distributed across parameters.

**Table 5.** Simulation-based required sample size (predictor distribution: Normal) by signal density, prevalence, and target AUC. Targets were 𝔼 (*b*_*n*_) ≥ 0.90 and Pr(*b*_*n*_ < 0.80) ≤ 0.20.

| $\phi_{\text{target}}$ | **AUC**<sub>target</sub> | **Signal** | $n_{\text{req}}$ | $\mathbb{E}(b_n)$ (95% CI) | $\Pr(b_n < 0.80)$ |
| --- | --- | --- | --- | --- | --- |
| 0.10 | 0.70 | Dense | 1746 | 0.910 (0.903, 0.918) | 11.5% |
|  |  | Sparse | 1746 | 0.902 (0.897, 0.907) | 14.3% |
|  | 0.80 | Dense | 904 | 0.903 (0.898, 0.908) | 13.8% |
|  |  | Sparse | 904 | 0.901 (0.896, 0.906) | 15.9% |
|  | 0.90 | Dense | 693 | 0.900 (0.895, 0.905) | 15.1% |
|  |  | Sparse | 712 | 0.901 (0.896, 0.906) | 16.1% |
| 0.20 | 0.70 | Dense | 1037 | 0.901 (0.896, 0.906) | 14.7% |
|  |  | Sparse | 1037 | 0.903 (0.897, 0.908) | 13.5% |
|  | 0.80 | Dense | 559 | 0.902 (0.897, 0.907) | 15.7% |
|  |  | Sparse | 598 | 0.901 (0.896, 0.906) | 16.3% |
|  | 0.90 | Dense | 482 | 0.900 (0.895, 0.905) | 14.7% |
|  |  | Sparse | 501 | 0.902 (0.897, 0.906) | 14.9% |

### Practical guidance: applying the findings in planning and reporting

The results suggest that, when the anticipated discrimination is high, development sample size planning should not rely on the closed-form recommendation alone. In such settings, we recommend using the Riley/`pmsampsize` calculation as a principled starting point, followed by a targeted stress test designed to detect whether the regularity conditions underpinning likelihood-based shrinkage approximations are plausible for the intended design.

In practice, the first step is to compute the benchmark *n*_Riley_ using `pmsampsize` with the prespecified number of candidate predictor parameters and the desired global shrinkage target (e.g., *S* ≥ 0.90), alongside the investigator’s anticipated performance inputs. This provides a transparent, reproducible baseline that is consistent with current guidance and is readily reportable in protocols. The second step is to conduct a small, scenario-matched simulation check at *n* = *n*_Riley_ that mirrors the intended modeling strategy and case-mix as closely as feasible. The objective of this check is not to exhaustively re-estimate *n*_req_, but to evaluate calibration-slope stability and to quantify the frequency of numerical irregularity indicators that can invalidate likelihood-based approximations. A practical implementation is to generate a moderate number of development replicates (e.g., several hundred) under a calibrated data-generating mechanism, achieving the anticipated discrimination, fit the planned model, and evaluate the truth-based logit-scale calibration slope on a large validation covariate set. If the resulting estimates indicate that 𝔼 (*b*) falls materially below the intended target or that Pr(*b* < 0.80) is unacceptably large, then *n*_Riley_ should be treated as optimistic for that design and the sample size should be increased or the modeling strategy strengthened.

The third step is to align the estimation and reporting strategy with the anticipated high-discrimination regime. If diagnostics indicate frequent separation-like behavior or extreme fitted risks at *n*_Riley_, investigators should consider estimation approaches that are more stable than unpenalized maximum likelihood, such as ridge penalization or other shrinkage methods, and should ensure that internal validation resamples the full modeling pipeline (including tuning) to provide optimism-corrected calibration summaries.

To operationalize these recommendations, Fig 4 in the Supplementary Material provides a decision flowchart that begins with the `pmsampsize` calculation, proceeds to a focused simulation-based stress test at *n*_Riley_, and then branches to either accept the benchmark sample size or revise the plan by increasing *n* and/or adopting a more stable estimation strategy. The flowchart is intended for direct use in protocols and statistical analysis plans, and it clarifies what quantities should be checked, what diagnostic thresholds should be reported, and how these checks connect to the calibration-stability criteria used throughout this study.

**Fig 4.**
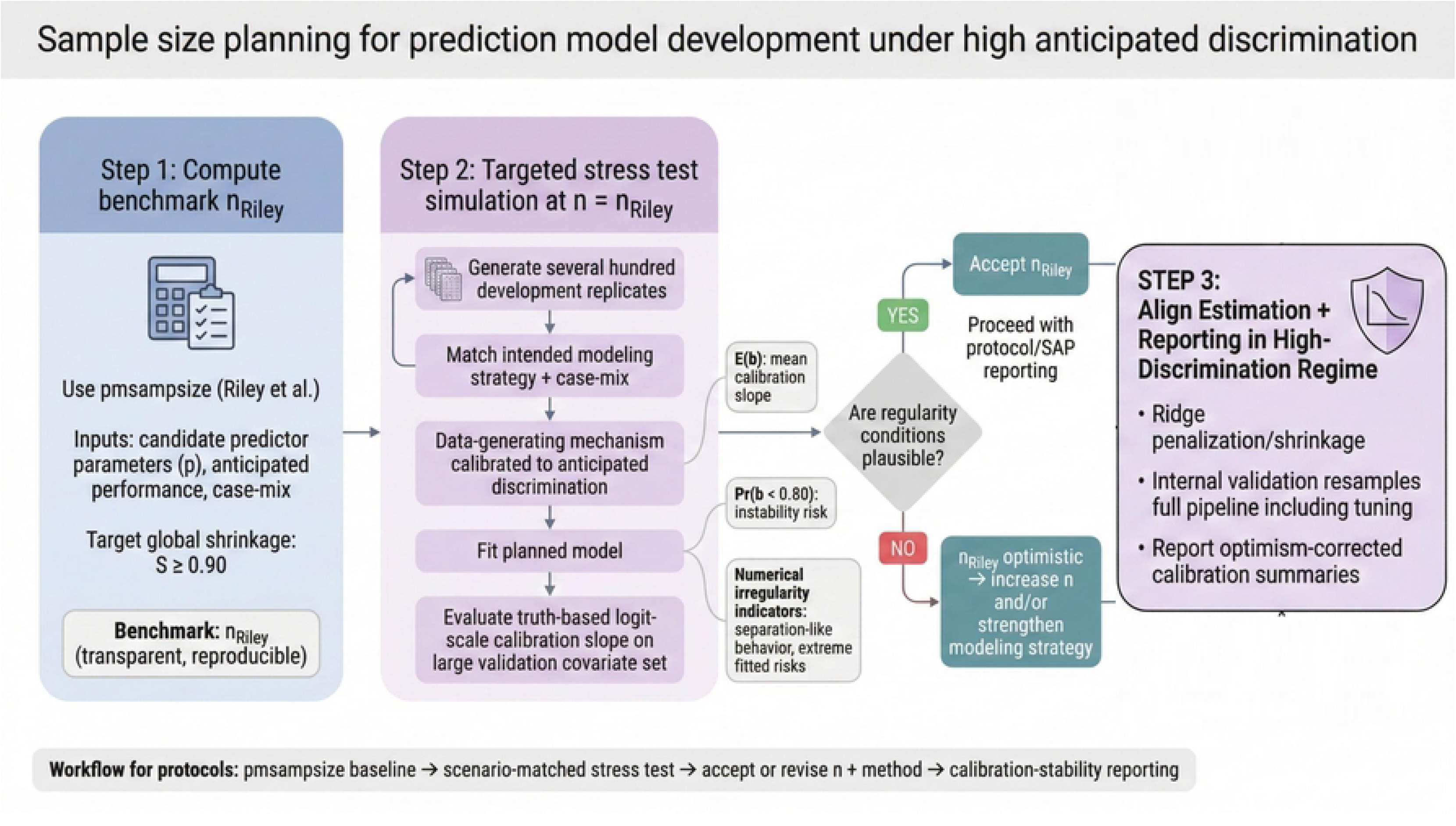
Practical guidance flowchart for sample size planning.

### Applied illustration

#### Dataset and candidate predictors

We provide a brief applied illustration using the stroke prediction dataset (open-sourced from Kaggle) [25], a retrospective cohort used to predict the likelihood of a patient suffering a stroke. Candidate predictors were prespecified based on clinical plausibility and routine availability at the point of care. The predictor set comprised three continuous variables (age, average glucose level, and body mass index) and seven categorical variables (gender, hypertension, heart disease, ever married, work type, residence type, and smoking status). The binary outcome *Y* indicates the occurrence of a stroke (*Y* = 1) versus no stroke (*Y* = 0).

Missing values in body mass index (BMI) were imputed using Multiple Imputation by Chained Equations (MICE) prior to analysis to ensure a complete dataset for ground-truth generation. Categorical predictors were represented using indicator (dummy) variables. After coding, the model included *p* = 15 predictor parameters, excluding the intercept.

For this illustration, we utilized the “Riley method” (implemented via `pmsampsize`) to calculate the minimum sample size required to minimize overfitting (targeting a shrinkage of < 10%). Based on a global population AUC of 0.847, the number of predictor parameter of 15 and a prevalence of 4.87%, the required sample size was calculated as *N* = 1492. Consequently, a training set of *N* = 1492 patients was sampled from the full cohort to simulate a realistic scenario of model development under sample size constraints.

The purpose of this section is descriptive and methodological: it illustrates how the calibration-focused evaluation framework used in the simulation study can be reported in an applied setting, rather than to draw etiologic conclusions about individual predictors.

#### Model specification and estimation

Let *Y*_*i*_ ∈ {0, 1} denote infection status for patient *i* and let **x**_*i*_ ∈ ℝ^*p*^ denote the corresponding vector of coded predictor values, where *p* is the number of estimated predictor parameters after dummy-variable expansion and excluding the intercept. We considered two logistic regression specifications with the same linear predictor structure but different estimation strategies. In both cases, the conditional event probability is modeled as

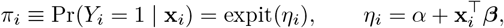

where *α* ∈ ℝ is the intercept and ***β*** ∈ ℝ^*p*^ is the vector of slope coefficients.

The first model used unpenalized maximum likelihood estimation (standard logistic regression). The estimator 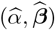 maximizes the binomial log-likelihood

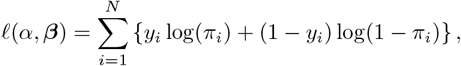

equivalently minimizing the negative log-likelihood. This model provides a conventional baseline against which penalized estimation can be compared under internal validation.

The second model used ridge-penalized logistic regression to stabilize estimation in the presence of limited effective information per parameter. Ridge estimation maximizes the penalized log-likelihood

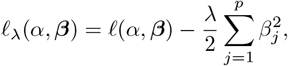

or, equivalently, minimizes

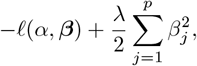

where *λ* ≥ 0 controls the degree of shrinkage applied to the slope coefficients; the intercept is not penalized. Continuous predictors were standardized for penalized estimation to ensure that the penalty operates comparably across coefficients, while indicator variables retained their natural coding. The ridge penalty parameter *λ* was selected by *K*-fold cross-validation using the binomial deviance as the objective, and the final penalized model was refit on the full dataset at the selected *λ*.

The model dimension was determined after preprocessing and coding. With 10 original candidate predictors (3 continuous and 7 categorical) and dummy-variable expansion for multi-level factors, the fitted models contained *p* = 15 slope parameters (excluding the intercept). The observed number of events was 80, yielding an events-per-parameter ratio of 80/15 ≈ 5.3. This quantity is reported descriptively to characterize the information available per parameter under the chosen specification; it is not used as a decision rule for adequacy, and it does not by itself determine expected predictive performance. The subsequent internal validation focuses directly on predictive calibration and discrimination rather than on heuristic thresholds.

#### Internal validation

Internal validation was performed using bootstrap optimism correction to quantify and adjust for over-optimism in apparent model performance when evaluation is conducted in the same dataset used for model development. The bootstrap procedure was applied separately to the unpenalized logistic regression model and to the ridge-penalized model, and in both cases it was applied to the full modeling pipeline as implemented. For the unpenalized model, the pipeline comprised predictor coding and model fitting by maximum likelihood. For the ridge model, the pipeline comprised predictor coding, standardization of continuous predictors, ridge model fitting, and selection of the penalty parameter *λ* by *K*-fold cross-validation within each bootstrap resample, followed by refitting the ridge model at the selected *λ*.

Let 𝒟 = {(*y*_*i*_, **x**_*i*_) : *i* = 1, …, *N*} denote the original dataset and let 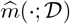 denote a fitted prediction model obtained by applying the relevant pipeline to 𝒟. The fitted model yields a linear predictor 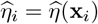 and predicted probability 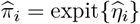 for each individual in any evaluation dataset. Apparent performance was computed by evaluating each metric in 𝒟 using predictions from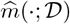 .

For *b* = 1, …, *B* bootstrap resamples, a bootstrap dataset 𝒟 ^(*b*)^ of size *N* was constructed by sampling with replacement from 𝒟. The complete modeling pipeline was then re-applied to 𝒟 ^(*b*)^ to obtain a bootstrap-fitted model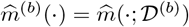. Each performance metric was evaluated twice for the same bootstrap-fitted model: first in the bootstrap sample 𝒟 ^(*b*)^ (apparent-in-bootstrap performance) and second in the original dataset 𝒟 (test-on-original performance). For a generic metric *T* (·), the bootstrap optimism in replicate *b* was defined as

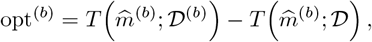

and the estimated optimism was 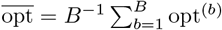. The optimism-corrected estimate was then obtained as

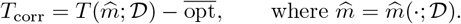

This procedure yields an internally validated estimate of expected performance in new individuals from the same target population under the modeling strategy considered.

Calibration was summarized on the logit scale by the calibration intercept and calibration slope as defined in Subsubsection Primary estimand: calibration slope in Section Methods. Discrimination was summarized by the area under the receiver operating characteristic curve (AUC) computed from predicted probabilities. All reported validation metrics in the applied illustration are bootstrap optimism-corrected unless stated otherwise.

### Findings and interpretation

Table 6 compares the bootstrap optimism-corrected performance of the unpenalized and ridge-penalized models. The unpenalized model exhibited signs of overfitting, with an optimism-corrected calibration slope of *E* (*b*_*n*_) = 0.802. This indicates that the fitted linear predictor varies too strongly, producing overly extreme risk predictions: a known instability of maximum-likelihood estimation when effective sample size is limited.

**Table 6.** Comparison of bootstrap optimism-corrected validation metrics: standard logistic regression versus ridge-penalized logistic regression. Calibration slope and calibration intercept are defined on the logit scale.

| Metric | Standard GLM | Ridge Regression |
| --- | --- | --- |
| $\mathbb{E}(b_n)$ | 0.802 | 1.013 |
| $\mathbb{E}(a_n)$ | -0.443 | 0.045 |
| $\Pr(b_n < 0.8)$ | 48.9% | 0.9% |
| Optimism-corrected AUC | 0.796 | 0.794 |

Ridge penalization successfully mitigated this instability. The probability of severe overfitting (defined as Pr(*b*_*n*_ < 0.8)) dropped from 48.9% in the unpenalized model to just 0.9% under ridge penalization. However, the ridge model yielded a slope above one (*E*(*b*_*n*_) = 1.013). This should not be interpreted as superior calibration; rather, it suggests a degree of underfitting or over-penalization, where the linear predictor is shrunk too aggressively, resulting in risk predictions that are insufficiently extreme.

Despite marked differences in calibration, discrimination remained comparable between the two strategies. The optimism-corrected AUC was 0.796 for the unpenalized model and 0.794 for the ridge model. This finding highlights that penalization primarily impacts the calibration (via shrinkage of the linear predictor) rather than the rank ordering of predicted risks.

Two key interpretations emerge from this application. First, predictor correlations in this real-world dataset likely reduced the effective information content, causing the standard sample size formula to be overly optimistic. Second, while penalization improves calibration stability, the shift from a slope of < 1 to > 1 illustrates that ridge regression requires careful tuning to avoid overly conservative shrinkage.

## Discussion

### Principal findings

This study evaluated the operating characteristics of the closed-form minimum development sample size criteria of Riley et al., as implemented in `pmsampsize`, against an empirical definition of the required development sample size based on logit-scale calibration stability under repeated development sampling. Across a factorial grid of predictor distributions and signal-density structures with a fixed number of candidate parameters (*p* = 10), the dominant empirical pattern was a discrimination-dependent divergence between the formula-based benchmark *n*_Riley_ and the simulation-based requirement *n*_req_. At moderate discrimination, the benchmark aligned closely with the required sample size, with small relative differences between *n*_Riley_ and *n*_req_. In contrast, as target discrimination increased into the high range, the benchmark increasingly recommended smaller development samples than were required to satisfy the prespecified calibration-stability criteria.

The divergence was systematic in the sense that the magnitude of underestimation grew as AUC_target_ increased. Although both approaches implied that fewer observations were needed as discrimination increased, the closed-form benchmark decreased more rapidly than the empirically required sample size for controlling calibration slope behavior, producing an increasing gap *n*_req_ − *n*_Riley_ at high AUC_target_. Mechanism diagnostics evaluated at *n* = *n*_Riley_ further indicated that, despite nominal convergence of unpenalized maximum-likelihood fits, separation-like irregularity and extreme fitted-risk behavior became common as discrimination increased. These findings are consistent with the interpretation that the likelihood-based information quantities underpinning the shrinkage approximation can be strained in high-discrimination regimes, yielding overly optimistic implied shrinkage and, consequently, overly small recommended development sample sizes.

Taken together, the principal contribution is empirical evidence that the accuracy of formula-based minimum sample size recommendations is primarily a function of discrimination in the settings examined here. This discrimination dependence provides a coherent explanation for why agreement with the empirical calibration-stability requirement is acceptable at moderate AUC_target_ yet degrades as AUC_target_ becomes high, motivating caution when applying closed-form criteria in high-discrimination development problems and supporting the use of complementary simulation-based assessment when such regimes are anticipated.

### Mechanistic interpretation

The discrimination-dependent discrepancy between *n*_req_ and *n*_Riley_ is most plausibly interpreted as a breakdown of the regularity conditions under which likelihood-based shrinkage approximations behave as intended. The Riley framework links the target global shrinkage factor to an information measure derived from the fitted model, commonly expressed through likelihood-ratio quantities such as Δχ^2^ (or equivalently a model fit measure related to 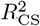). These relationships are justified under standard maximum-likelihood theory for logistic regression, which assumes that the maximum-likelihood estimator exists, that the log-likelihood is well approximated by a quadratic form in a neighborhood of the optimum, and that the curvature (Fisher information) provides a stable characterization of uncertainty. When these conditions hold, the degree of overfitting in the fitted linear predictor can be summarized by a single multiplicative shrinkage factor, and the resulting calibration slope in new data is expected to be close to that factor in repeated sampling.

Our mechanism diagnostics indicate that, as discrimination increases, development-data fits increasingly enter regimes that are consistent with quasi-separation, even when the fitting routine reports nominal convergence. In such replicates, the fitted linear predictor takes very large absolute values for at least some individuals, and the corresponding fitted probabilities become numerically extreme. This separation-like behavior does not necessarily manifest as non-convergence or as obviously divergent coefficient estimates at a fixed coefficient-magnitude cutoff; rather, it appears primarily through extreme fitted risks and large log-odds for a subset of observations. From the standpoint of likelihood theory, these regimes are consequential because the Bernoulli log-likelihood becomes dominated by observations whose fitted probabilities are very close to 0 or 1, and the effective curvature of the log-likelihood can become highly uneven across directions in parameter space. Consequently, likelihood-based summary quantities such as Δχ^2^ can be inflated or otherwise perturbed relative to what would be expected under well-behaved sampling distributions, because the apparent separation leads to very large improvements in the fitted likelihood even when the fitted linear predictor is unstable from a predictive perspective. If Δχ^2^ is inflated at a given *n*, then shrinkage approximations of the form 1 − *p*/Δχ^2^ will tend to be overly optimistic, implying a larger expected shrinkage (and hence recommending a smaller *n*) than is warranted to achieve stable out-of-sample calibration.

This interpretation is consistent with the empirical pattern that *n*_Riley_ decreases sharply as AUC_target_ becomes high, whereas *n*_req_ decreases more gradually when defined by repeated-sampling constraints on the logit-scale calibration slope. The truth-based calibration slope estimand used in the simulation study is designed to isolate instability attributable to the fitted linear predictor itself by evaluating calibration against the known true log-odds on a large validation covariate set. The fact that this estimand remains numerically defined in the scenarios examined, even when separation warnings and extreme fitted risks become common, indicates that the principal issue is not failure of the calibration-slope estimand but rather instability of the development fit that compromises the validity of likelihood-based approximations. Overall, the mechanism results support a coherent account: as discrimination increases, separation-like behavior becomes frequent enough that likelihood-based information measures cease to provide a reliable proxy for the effective overfitting of the fitted linear predictor, leading the closed-form criteria to understate the development sample size required for calibration stability under repeated sampling.

### Relation to prior work

Our findings sit within a rapidly developing literature on performance-based sample size planning for prediction model development and provide targeted evidence about when closed-form criteria may become unreliable. Riley and colleagues formalized minimum development sample size criteria for logistic prediction models by linking prespecified targets for overfitting control and model-fit optimism to quantities such as the global shrinkage factor and the Cox–Snell *R*^2^ (and its associated maximum), yielding an implementable set of closed-form calculations that have been disseminated widely through `pmsampsize`. This framework has substantially advanced practice beyond historical EPV heuristics by making explicit the dependence of required sample size on anticipated model performance and by emphasizing calibration-oriented objectives rather than solely parameter estimation precision.

At the same time, prior methodological work has emphasized that small-sample behavior of logistic regression can be problematic in settings that challenge the assumptions underpinning asymptotic likelihood theory. Earlier studies of overfitting and shrinkage in generalized linear models, including foundational work on heuristic and formal shrinkage approaches, highlighted that the stability of the fitted linear predictor is central for out-of-sample calibration. More recent commentary by van Smeden and colleagues has reinforced that sample size requirements for prediction cannot be reduced to a single rule and may depend strongly on design features and on how model performance targets are operationalized. Our results align with these general messages while adding a specific, discrimination-focused characterization of where and how a widely used closed-form method can become optimistic.

The principal contribution of the present study, relative to existing work, is twofold. First, we provide a direct empirical mapping between target discrimination and the magnitude of divergence between the closed-form benchmark *n*_Riley_ and a simulation-defined required sample size *n*_req_ based on repeated-sampling behavior of a clearly defined, logit-scale calibration slope estimand. This places the comparison on an estimand that is explicitly aligned with calibration and is interpretable in relation to the shrinkage target used by the Riley criteria. Second, we complement this primary comparison with a mechanism-oriented diagnostic layer that quantifies separation-like irregularity at *n* = *n*_Riley_ and shows that such irregularity increases sharply as discrimination increases, even in the absence of overt optimizer failure. This diagnostic mapping provides concrete evidence for a plausible mechanism: likelihood-based quantities used in shrinkage approximations can be perturbed in high-discrimination regimes because the fitted likelihood becomes dominated by near-deterministic predictions, weakening the connection between Δχ^2^-based information measures and the effective overfitting of the linear predictor.

In this sense, our study extends prior work by moving beyond documenting discrepancies toward explaining them using operational diagnostics that can be implemented routinely. The implication is not that performance-based criteria are unhelpful, but that their reliability depends on the extent to which regularity conditions hold in the intended operating regime. By identifying discrimination as a primary axis along which these conditions can be strained, and by providing empirically grounded diagnostics to detect such strain, our results offer an actionable refinement to current guidance for applying `pmsampsize` in contemporary prediction model development studies.

### Implications for practice

The results have practical implications for planning and analysing prediction model development studies that target strong discrimination. In settings where anticipated discrimination is moderate, the closed-form criteria implemented in pmsampsize provided sample size recommendations that were broadly consistent with the simulation-defined requirement based on repeated-sampling calibration-slope behavior. In such scenarios, `pmsampsize` remains a defensible and efficient first-line tool for prospective planning, provided that inputs are specified transparently and that the resulting recommended sample size is interpreted as a minimum under the assumed operating conditions. In particular, when the objective is to limit overfitting of an unpenalized logistic model and to align development planning with an explicit shrinkage target, the `pmsampsize` framework offers a principled alternative to EPV heuristics and encourages calibration-focused thinking.

When anticipated discrimination is high, however, our findings indicate that reliance on the closed-form benchmark alone may be optimistic with respect to calibration stability under repeated development sampling. The practical consequence is that a study planned solely on *n*_Riley_ may yield models whose fitted linear predictors are more variable than intended, even if nominal convergence is achieved, with an increased probability of extreme risk predictions and separation-like behavior. For applied researchers, this motivates augmenting formula-based planning with additional safeguards. One pragmatic approach is to treat *n*_Riley_ as a lower bound and to perform a scenario-specific simulation study that mirrors the planned modeling strategy and the anticipated distribution of covariates, using clearly stated calibration targets and estimands. Such simulation-based planning is particularly informative when development is expected to occur in regimes where numerical regularity may be strained, because it evaluates the operating characteristics of the complete analysis pipeline directly rather than relying on asymptotic approximations.

A second implication is that the modeling strategy used in development should be aligned with the sample size regime. If penalization is planned a priori, then sample size justification should not be based exclusively on criteria derived for maximum-likelihood estimation. Penalized regression, including ridge-type shrinkage, can mitigate instability of coefficient estimates and reduce tail-risk of severe miscalibration, but it introduces its own tuning choices that can shift calibration slope above or below 1 depending on penalty strength. Accordingly, when penalization is used, researchers should report the penalty selection approach, present optimism-corrected calibration intercept and slope on the logit scale, and include a calibration curve to evaluate calibration beyond a single summary statistic. Penalization should be framed as a modeling choice that trades variance reduction against potential underfitting, rather than as a post hoc remedy for small samples.

More generally, we recommend an integrated workflow for high-discrimination development studies. Formula-based calculations can be used to establish an initial minimum sample size target and to structure reporting of assumptions. This should be complemented by either simulation-based evaluation of calibration stability under the anticipated regime or, where this is infeasible, by prespecification of a penalized modeling strategy with rigorous internal validation and transparent reporting of calibration. In all cases, researchers should anticipate that high discrimination can coincide with separation-like behavior in unpenalized logistic regression and should plan diagnostics accordingly, including routine checks for extreme fitted risks and sensitivity of calibration estimates. These steps provide a practical route to maintaining interpretability and calibration reliability while retaining the convenience and transparency of closed-form planning tools.

### Strengths

This study has several methodological strengths that support the credibility and interpretability of its findings. First, the simulation framework was constructed under a known and correctly specified data-generating mechanism, so that the “truth” underlying each scenario was fully defined. This eliminates ambiguity about the target of inference and avoids conflating model misspecification with sample size effects, thereby enabling a clean evaluation of how development sample size interacts with discrimination to influence calibration stability.

Second, the study was anchored to an explicit, prespecified estimand for calibration performance. The required development sample size was defined in terms of repeated-sampling behavior of an out-of-sample calibration slope on the logit scale, with targets on both the mean slope and the lower-tail probability of severe miscalibration. This estimand-based design aligns directly with the practical goal of controlling overfitting and yields results that can be interpreted without relying on informal heuristics. In addition, by defining performance targets at the population level and evaluating them via repeated development sampling, the study avoids interpretations that depend on idiosyncratic properties of any single development dataset.

Third, calibration slope estimation was implemented using a large, fixed validation covariate set with deterministic evaluation of the true event probabilities implied by the data-generating mechanism. This design choice effectively removes outcome noise from validation, yielding a high-precision estimate of the logit-scale calibration relationship between the fitted linear predictor and the true log-odds. As a result, Monte Carlo variability in the reported calibration slopes is dominated by variation in the development fit, which is the relevant source of uncertainty for development sample size planning.

Fourth, the algorithms used to identify *n*_req_ were designed for computational efficiency and reproducibility while maintaining conservative decision-making. The bisection search reduced the number of evaluated sample sizes relative to exhaustive grid approaches, replication was adaptively increased where needed, and decision rules were based on Monte Carlo uncertainty bounds rather than dichotomous hypothesis testing. Parallel computation was coupled with reproducible random number generation, and scenario-specific computations were structured to enable caching and consistent reuse of validation covariates. Together, these features strengthen the reliability of the reported comparisons and facilitate transparent replication and extension of the study by other investigators.

### Limitations

Several limitations should be considered when interpreting the generality of these findings. First, the simulation design fixed the number of candidate predictor parameters at *p* = 10 and used a small set of prespecified prevalence strata and discrimination targets. Although this choice supports a controlled assessment of how discrepancies evolve as a function of discrimination, it does not establish that the same magnitude of divergence will occur for substantially larger *p*, or for more finely resolved or more extreme discrimination regimes. In particular, the behavior of likelihood-based approximations and the frequency of separation-like irregularities may differ when *p* is larger relative to the number of events, even if the target AUC is held constant.

Second, predictors were generated under a simplifying independence assumption within each data-generating mechanism. This was intentional to isolate the role of discrimination from confounding effects of correlation structure, but it limits direct extrapolation to applied settings where predictor correlation is common and can materially affect both discrimination and estimation stability. The results therefore should not be interpreted as establishing that discrimination is the only driver of formula performance; rather, they demonstrate a clear discrimination-dependent pattern under a controlled covariance structure. Extending the design to correlated predictors, including both mild and strong correlation, would be necessary to quantify how correlation interacts with discrimination in determining the accuracy of closed-form sample size approximations.

Third, although three distinct predictor distribution families were examined (normal, standardized skewed continuous, and binary), these data-generating mechanisms remain stylized representations of clinical predictor sets. Real-world predictor distributions may exhibit heavier tails, mixed discrete–continuous structures, measurement error, and complex dependence patterns that are not captured by the present design. Similarly, the simulation grid for discrimination was restricted to AUC_target_ ∈ {0.70, 0.75, 0.80, 0.85, 0.90}, which provides coverage of commonly reported discrimination levels but does not explore very high discrimination beyond 0.90, where numerical irregularities may become even more prominent. A denser discrimination grid could also help to locate more precisely the range at which formula performance begins to degrade.

Fourth, the validation strategy for the truth-based calibration slope relied on a large, fixed covariate-only validation set generated from the same predictor distribution as the development samples. This approach was adopted to remove validation outcome noise and to target the logit-scale calibration relationship to the known data-generating mechanism, but it represents an idealized evaluation regime. In applied research, external validation uses realized outcomes and may involve shifts in case-mix or predictor distributions. Moreover, the use of a fixed validation covariate set reduces Monte Carlo variability and improves precision, but it also conditions the evaluation on a particular realization of covariates; while N_val_ was chosen to be sufficiently large to make this dependence negligible in practice, alternative designs that regenerate validation covariates per replicate could be used to confirm robustness of the reported patterns.

Finally, several modeling complexities that commonly arise in practice were not addressed. The development model was correctly specified and linear on the logit scale, with no interactions, nonlinear effects, time-to-event structure, or variable selection. The study also did not incorporate missing data mechanisms, imputation, measurement error, or informative censoring. Each of these features can affect effective model complexity, estimation stability, and calibration performance, and may therefore influence both simulation-based and formula-based sample size requirements. Consequently, the present results should be interpreted as characterizing the operating behavior of the Riley/`pmsampsize` framework under controlled, correctly specified logistic models, and as motivating further work to map discrimination-dependent discrepancies under additional sources of complexity that are typical of applied prediction modeling.

### Future work

Several extensions would strengthen the practical scope of the present findings and help translate them into more general guidance for study planning. A priority is to expand the simulation design to include realistic predictor correlation structures. Future work should consider both exchangeable and heterogeneous correlation patterns and should explicitly evaluate how correlation interacts with discrimination to influence the accuracy of likelihood-based shrinkage approximations and the frequency of separation-like irregularities at formula-recommended sample sizes. Similarly, extending the evaluation to higher-dimensional settings, with substantially larger numbers of candidate parameters *p*, would clarify how discrimination-dependent discrepancies scale as the events-per-parameter ratio decreases and as the curvature assumptions underlying closed-form criteria become more fragile.

An important methodological extension is to adapt the framework to time-to-event prediction models. Riley-type sample size criteria have been developed for Cox and related models, but high-discrimination behavior and numerical irregularities in survival settings may differ due to censoring, risk set structure, and time-varying baseline hazards. Extending the present simulation-based definition of required development sample size to survival outcomes, using appropriately defined calibration estimands and validation schemes, would permit a parallel investigation of whether discrimination-driven divergence arises in those settings and whether the mechanism is analogous.

Finally, future work should evaluate alternative estimators within the planning loop rather than treating maximum-likelihood logistic regression as the default development method. In particular, bias-reduced and separation-robust approaches such as Firth’s penalized likelihood, Bayesian logistic regression with weakly informative priors, and other penalized likelihood estimators could be incorporated directly into the simulation-based determination of *n*_req_. This would allow the required sample size definition to be aligned with the intended estimation strategy, and would quantify whether the discrimination-dependent discrepancy is attenuated when the development estimator regularizes or stabilizes the likelihood in regimes where standard maximum likelihood is prone to separation-like behavior. Such work would also support practice-oriented recommendations that jointly specify an estimation strategy and a sample size target, rather than treating sample size planning and estimation as separable decisions.

## Conclusions

In this study we evaluated the closed-form minimum development sample size criteria of Riley et al., as implemented in `pmsampsize`, against a simulation-based definition of the required sample size that targets logit-scale calibration slope stability under repeated development sampling. Across the factorial scenarios considered, the formula-based benchmark provided a close approximation to the simulation-based requirement at moderate discrimination, but became progressively optimistic as the target AUC increased, with underestimation of the required development sample size growing monotonically in high-discrimination regimes. Mechanism diagnostics at *n* = *n*_Riley_ showed that separation-like numerical irregularity, expressed primarily as extreme fitted risks and frequent separation warnings despite nominal optimizer convergence, increased sharply with discrimination and provides a plausible explanation for the observed deterioration of likelihood-based shrinkage approximations in these settings. For applied planning, these results support continued use of `pmsampsize` as a reasonable baseline when anticipated discrimination is moderate, but suggest that high-discrimination applications should be accompanied by explicit numerical diagnostics and, where feasible, augmented by simulation-based planning aligned to the intended estimand and modeling strategy; in settings where instability is anticipated, penalized or bias-reduced estimation should be considered as part of a principled development and validation workflow.

## Data Availability

All relevant data are within the manuscript and its Supporting Information files.

## Notes

### Competing Interest Statement

The authors have declared no competing interest.

### Funding Statement

The author(s) received no specific funding for this work.

